# A Poisson Process Life Expectancy framework for optimising patient lifetime during chemotherapy

**DOI:** 10.64898/2026.06.15.26354436

**Authors:** Byron D. E. Tzamarias, Nigel J. Burroughs

## Abstract

Cancer therapy must balance between two competing objectives - treatment efficacy against the tumour and the risk of treatment related severe adverse events, including patient death. While optimal control theory (OCT) provides a principled framework for designing dosing protocols, most existing formulations rely on optimising heuristic cost functionals that lack direct clinical interpretability. In clinical practice treatment efficacy and patient tolerability are primarily assessed through survival metrics and adverse event rates. Here we introduce the Continuous Lifetime Payoff (CLP), a novel OCT objective functional that directly links treatment decisions to patient survival. It explicitly incorporates tumour dynamics, tumour eradication, and patient mortality from tumour progression, drug-related toxicity and age. Mortality is modelled as a Poisson processes, a framework that enables the patient’s expected lifetime to be computed. The model and objective are biologically interpretable and can in principle be calibrated using clinical data. We demonstrate the approach for a logistic tumour growth model, illustrating the trade-offs between tumour control and toxicity. We fit age-related mortality from national life tables and demonstrate parameter inferability from overall survival curves using simulated data. The CLP offers a clinically grounded OCT objective and provides a foundation for data-driven optimisation of chemotherapy regimens.

## 1 Introduction

Applying mathematical optimisation methods to cancer treatment requires quantifiable measures of the efficacy of treatment against the tumour and the impact of treatment on the patient. This is extremely challenging as there are no obvious candidates, whilst there are many ways to interpret efficacy of treatment and the negative effects of treatment are multi-faceted. Hence heuristic measures have typically been used that represent desirable objectives of treatment, for instance minimising both the final tumour size and the amount of treatment. Optimising cancer therapy is thus a multi-objective optimisation problem. This multi-objective optimisation problem can be solved within the context of tumour response dynamics by constructing a weighted sum of the objectives, so called scalarisation, [19], giving a classical optimal control theory (OCT) problem, [61]. Unfortunately, scalarisation raises another issue - evaluating the net benefit of a therapy to the patient as a weighted sum of *“*harm and benefit*”* objectives, often with different units, has no medical justification. Here we develop an event based framework, incorporating patient death and tumour elimination events, that quantitatively calibrates the impact of treatment through an increase/decrease of these event rates. The net benefit of treatment to a patient is then naturally quantified by the expected extended lifetime, giving an objective that can be used to determine the optimal therapy within an OCT framework. This also provides a mapping to clinical outcomes and survival distributions, enabling model parameters to be fitted from clinical data. Our framework thus solves 2 major problems in cancer therapy optimisation: the problem of quantifying the efficacy and (negative) impacts of a therapy, and combining these positive and negative impacts within a single measure – the expected patient lifetime.

Mathematically, OCT determines the time dependence of a control (here the drug concentration over time) that optimises a specified objective, [20, 31, 61]. In applications to cancer therapy, the objective quantifies the net benefit (or cost) of treatment to the patient, a functional of the tumour dynamics (usually described by a system of ordinary differential equations) and the control over a finite, predetermined time interval [0, *T*], where *T* is referred to as the time horizon. By framing treatment planning as an optimisation problem, OCT provides a principled alternative to subjective approaches, grounded in rigorous control and optimisation theory, and has been applied to a wide array of cancer therapy optimisation problems [31, 39, 58, 61, 64]. In most applications of OCT to cancer therapy a weighted sum of competing objectives is used, for instance the sum of the final tumour size at the horizon time *T* to gauge impact on the tumour and the total drug administered over the duration [0, *T*] as a proxy for patient toxicity [31], both of which need to be minimised. This formulation presents several issues: each objective is only represented as a qualitative penalty term, lacking direct medical or clinical relevance. In particular, drug toxicity penalty terms are often chosen for mathematical convenience (such as the total squared drug dose (*L*_2_ norm) that ensures problem convexity and thus guarantees unique optimal solutions [61]) rather than reflecting measurable biological or clinical endpoints, such as minimising chemotherapy adverse events. Secondly, the weights used to trade off tumour control efficacy against toxicity to the patient have no biological justification and are inherently subjective. Changes in these weights can lead to qualitatively different optimal treatment strategies [20, 44]. Multi-objective optimal control (MOOC) addresses these issues by treating tumour control and toxicity as separate objectives and seek Pareto-optimal solutions, i.e., regimens for which no objective can be further improved without worsening another [22]. The resulting Pareto front provides a spectrum of non-dominated strategies, explicitly revealing trade-offs between competing goals and allowing exploration of multiple treatment options; however, selecting a single clinically relevant strategy remains subjective as preferences among Pareto-optimal solutions are rarely standardised or clinically explicit [27, 59].

Biologically grounded and clinically interpretable optimisation criteria that reflect realistic, patient-centred treatment goals remain largely unexplored. In clinical practice therapy outcomes are evaluated using criteria such as survival endpoints [15], for instance overall survival (OS), whilst tumour response is often classified as an overall, complete, or partial response. In addition treatment tolerability is assessed by the rate and severity of adverse events during treatment [38], a 5 point scale from mild (no interventions indicated) to death. Severe adverse effects (SAEs) can result in a treatment pause/cessation or dose reduction that all jeopardise patient survival, whilst in more severe cases can result in treatment-related mortality [38]. Patient lifetime provides a natural objective for treatment optimization that incorporates these clinical measures, since the consequences of drug toxicity (adverse effects) is balanced against control of the tumour through their impact on the patient’s lifetime. Metrics linked to survival have been used previously in optimal cancer therapy problems, for instance maximising the time that the tumour remains within an acceptable (safe) size range in patients with incurable cancer [7, 48, 73]. This safe zone is essentially a progression-free survival criteria. From a clinical perspective patient survival under chemotherapy is determined by discrete treatment-related events, such as severe adverse events (SAEs), emergence of secondary (metastatic) tumours and disease relapse. This viewpoint has motivated sequentially adaptive medical decision-making or dynamic treatment regime modelling [10]. These approaches formulate multi-stage therapy selection within a Markov decision-making framework and have been applied to optimise sequential drug use (e.g. in acute myeloid leukaemia [29]), but do not explicitly model tumour dynamics. Recent studies have developed mathematical frameworks that couple tumour growth dynamics with survival modelling to simulate clinical trial outcomes and generate survival curves [37, 74]. In [37] clinical trials were simulated to investigate chemotherapy combined with surgery for high-grade serous ovarian cancer, integrating branching process models of tumour growth with white blood cell dynamics to capture treatment-related toxicities such as anaemia and neutropenia. Survival was assessed using Kaplan–Meier estimators, with good agreement between model predictions and clinical data. In [74], a unified framework was proposed that combined ODE-based tumour dynamics with Poisson process models of metastasis and survival, demonstrating parameter identifiability and good agreement with simulated data. Neither these works explored optimal therapy.

In previous work, we proposed maximising the expected patient lifetime within an ODE modelling framework [8, 69]. We utilised the probability of tumour elimination (the tumour control probability, TCP) from branching processes, [79]; the expected lifetime being an average over the outcomes of tumour elimination and no elimination. In [8] we used an approximation for the elimination probability with an exponential dependence in the tumour size and discounted the expected lifetime by the detrimental effects of drug toxicity (interpretable as a measure of the quality of life, QoL). Bang-bang solutions with a single switch were derived using Pontryagin’s Maximum principle. We extended this analysis in [69], where we introduced the Life Expectancy Payoff (LEP) which maximises a patient’s expected lifetime averaged over key therapy events, including complete remission, treatment failure and SAEs. Event probabilities were evaluated analytically giving a closed form expression for the objective functional. We proved that optimal protocols were bang-bang using Pontryagin’s Maximum principle, in fact the only switching solution had a single switch from maximum tolerated dose to no treatment. The LEP, by maximising the patient’s expected lifetime averaged over all therapy outcomes, explicitly balances the competing goals of minimising drug toxicity and maximising tumour elimination in a biologically interpretable manner. However, the LEP has a notable limitation; effects of tumour size on patient mortality (and other event rates) could not be included (only the probability of tumour elimination was dependent on the tumour size history). Predetermined lifetimes were used for disease free individuals and patients with a tumour, cured patients acquiring the disease free lifetime. Yet, tumour size clearly impacts a patient’s lifetime and prognosis, with many studies reporting that larger tumour sizes are positively correlated with increased patient mortality, including breast cancer [24, 50, 71], lung cancer [51, 81], liver cancer [76] and pancreatic cancer [67]. Incorporating the effect of tumour progression directly on survival would therefore provide a more realistic and biologically justifiable formulation of the LEP.

Here we develop the Continuous Lifetime Payoff (CLP), an extension of the LEP that directly accounts for the time of tumour elimination. The payoff incorporates mortality related to tumour growth and severe chemotherapy adverse effects, in addition to baseline age-dependent mortality. Mortality causes are modelled as independent Poisson processes, the patient’s expected lifetime and outcome then following from the respective survival probabilities. Thus, the CLP is framed in terms of continuous stochastic processes, effectively capturing the direct contribution of key events to the patient’s lifetime as well as the likelihood of each event (and the associated outcome) occurring. There are no free parameters in our objective, *i*.*e*. the positive and negative effects of treatment are implicitly weighted by the impact of treatment on the rates of their respective events. We therefore do not have a multi-objective problem to solve as is typical of OCT applications to cancer that require a subjective weighting of the competing objectives [20, 61]. The proposed optimal control formulation is very general, the key ingredients being a tumour growth model and a set of key events that impact patient prognosis with tumour and drug dependent rates. All parameters thus have a direct biological interpretation. Consequently, the CLP can, in principle, be fully parameterised using real-world data, provided that suitable data are available. For instance, we use life-tables to callibrate age-related mortality [53].

This paper is organised as follows. In Section 2, the probability distribution for tumour elimination is introduced and in Section 3 a patient’s mortality probability is defined as a function of tumour size, administered drug dosage, and patient age. In Section 4 the CLP objective functional is developed. The expected lifetime under chemotherapy is decomposed into two cases: tumour elimination at a specific time and tumour persistence; these scenarios are then integrated to derive the overall expected lifetime, accounting for both successful elimination and treatment failure. In Section 5, a case study is presented where the objective functional is maximized under logistic tumour-growth. In section 6 we examine survival curves as a means of parameter estimation for our model. Finally, Section 7 offers concluding remarks.

## 2 Tumour elimination probabilities

To account for the impact of tumour elimination on patient survival, we need to determine the probability density function of the time of tumour elimination. In this section, we build on key concepts introduced in the LEP [69], using the tumour control probability (TCP) of branching processes to derive a distribution for elimination events occurring during or after treatment.

### 2.1 The Tumour Control Probability (TCP)

Consider a chemotherapy treatment that takes place within a time interval [0, *T*_0_], for some positive horizon time *T*_0_. We analyse optimal therapy for deterministic single-compartment tumour dynamic models of the form,

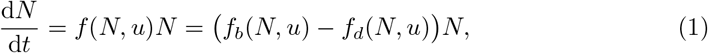

where *N*(*t*) is the tumour size and *u*(*t*) the concentration of drug administered at time *t*. Without loss of generality, *u*(*t*) ∈ [0, 1], where *u* = 1 is the maximum tolerated dose (MTD). The functions *f*_*b*_(*N, u*) and *f*_*d*_(*N, u*) represent the per-capita growth and death rates of cancer cells, respectively. The death rate is assumed to increase with both tumour size and drug concentration *u, i*.*e*. 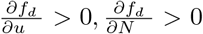. This includes natural death (in absence of drug), for instance due to resource limitation. In the absence of treatment (*u* = 0), the tumour grows and approaches a steady state at the carrying capacity *K*, given by *f*_*b*_(*K*, 0) = *f*_*d*_(*K*, 0). Under MTD drug administration (*u* = 1), we assume the tumour regresses, requiring that *f*_*b*_(*N*, 1) *< f*_*d*_(*N*, 1) for all *N* ∈ [0, *K*].

To derive an analytical form for the probability that the tumour is eliminated by a time *t* ∈ (0, ∞), we employ the tumour control probability (TCP) [79]. The TCP, defined as the probability that the tumour size is 0 at a time *t*, can be computed for time varying birth and death rates. We use birth and death rates *a*(*t*) = *f*_*b*_(*N*(*t*), *u*(*t*)) and *b*(*t*) = *f*_*d*_(*N*(*t*), *u*(*t*)) respectively, where *N*(*t*) satisfies (1), see [69]. Here we assume that for small tumour sizes the dependence on *N*(*t*) is negligible (competition effects only become important when cell numbers are large, in fact of the order of the capacity *K*), thus the branching process rates are then independent of the tumour size and the TCP can be computed exactly [79]. The nonlinear branching process with birth and death rates *f*_*b*_(*Z*_*t*_, *u*), *f*_*d*_(*Z*_*t*_, *u*), tumour cell count *Z*_*t*_ at time *t*, has mean dynamics given by (1) under the mean field closure approximation. The TCP, for large initial tumour size *N*_0_, is then well approximated by, [69],

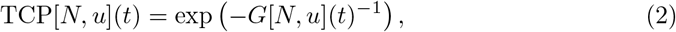

where

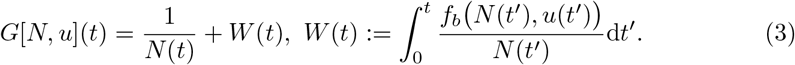

This is independent of *N*_0_. The notation TCP[*N, u*](*t*), signifies that the TCP is a functional of the tumour history *N*(*t*) and of the drug regimen *u*(*t*), while the argument (*t*) indicates the current time *t*. From the branching-process perspective, TCP[*N, u*](*t*) corresponds to the cumulative extinction probability up to time *t*, accounting for the full history of time-dependent birth and death rates that depend on tumour size and treatment. A straight forward calculation gives the time derivative,

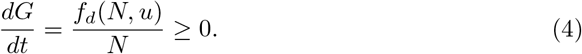

Thus, the TCP is non-decreasing as expected.

The TCP has two key properties. If the natural branching-process death rate is zero, (*f*_*d*_(*N*, 0) = 0), after treatment the TCP remains constant. Conversely, if *f*_*d*_(*K*, 0) *>* 0, the TCP increases monotonically and asymptotically approaches one as *t* → ∞ since stochastic fluctuations eventually eliminate the tumour. A proof is provided in Supplement A. Thus, rare stochastic extinction events driven by population fluctuations eventually eliminate the tumour. When the tumour stays near carrying capacity (i.e., *N*(*t*) ≥ *ζK*, with *ζ*1) on time scales shorter than a human lifespan, these stochastic fluctuations have a negligible impact on the TCP (see Supplement B) and can be ignored. This is also evident from Equation 4, giving 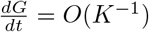. In contrast, when treatment reduces the tumour size to single digits, stochastic extinction events are more frequent; these time periods thus dominate the contributions to the elimination probability. Since we are only concerned with finite time scales relevant to patient life expectancy, we suppress die-out events whenever *N*(*t*) gets larger than a threshold *N*_*_ = *ζK* (with *ζ*1), regardless of treatment, and restore die-out dynamics if/whenever *N*(*t*) *< N*_*_. The TCP is given by Equation 2 with modified functional *G*[*N, u*](*t*),

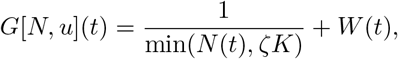

with *W* (*t*) satisfying

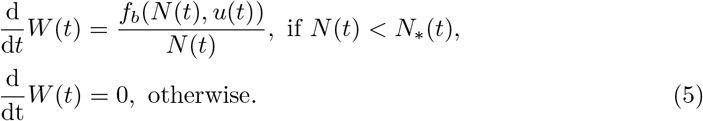

Thus,

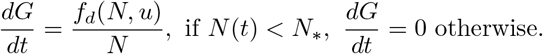

### 2.2 The Tumour Elimination Time Distribution

Our approximation (2), has a negligible but positive probability that the tumour is eliminated before therapy starts since TCP[*N, u*](0) *>* 0. We condition on the tumour existing at *t* = 0 by defining the cumulative elimination probability as

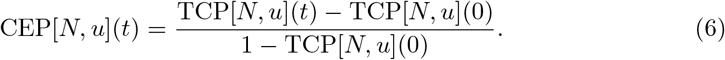

Since 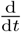 TCP[*N*, 0](*t*) *>* 0 provided *f*_*d*_(*N*, 0) *>* 0, spontaneous tumour elimination (without therapy) is theoretically possible. However, since tumours are typically detected with more than 10^6^ cells [2], this probability is negligible over a finite horizon time in the absence treatment.

Taking the limit of the CEP as *t* → ∞ the probability of tumour elimination (PTE) can be derived, i.e. the probability that the tumour is eventually eliminated,

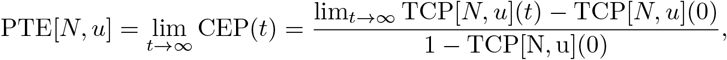

which is a function of the state trajectory *N*(*t*) and control history *u*(*t*). After therapy ends, (*t > T*_0_), the average tumour size *N* → *K*, (1), and thus there exists a time *t*_*_ = **max**{*t*|*N*(*t*) = *N*_*_}, ie the largest time *t*_*_ with the property *N*(*t*_*_) = *N*_*_. Then lim_*t*→∞_ TCP[*N, u*](*t*) = TCP[*N, u*](*t*_*_). It is convenient to introduce a time *T*_*ζ*_ *> T*_0_ such that the interval *T*_*ζ*_ − *T*_0_ is sufficiently long for the tumour to approach carrying capacity, i.e., *N*(*T*_*ζ*_) ≥ *ζK*, for any admissible regimen on [0, *T*_0_]. Then

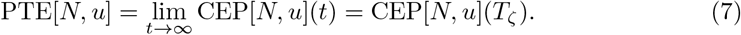

Importantly, the PTE resolves a key issue in OCT applications to cancer chemotherapy. It provides a robust measure of a treatment’s ability to control the tumour and is independent of the time horizon *T*_0_ if drug administration has ended by *T*_0_ since the trajectory (*N*(*t*), *u*(*t*)) is unchanged. This distinguishes the PTE from traditional OCT measures, such as tumour size at a fixed horizon *N*(*T*_0_) or the cumulative tumour burden 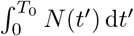, which are explicitly dependent on *T*_*0*_ and thus the optimal solution depends on *T*_0_.

To model the influence of tumour burden on patient mortality, the timing of tumour elimination must be taken into account; earlier elimination will be associated with improved survival. The tumour elimination time *τ* is a random variable with probability density function (PDF),

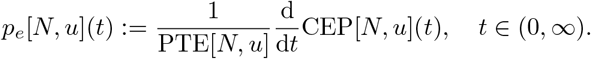

conditioned on elimination. Note that tumour elimination can occur after the horizon *T*_0_, for instance if *N*(*T*_0_) is small. However, because we removed stochastic fluctuations once *N > N*_*_ = *ζK*, elimination time is bounded by *t*_*_.

## 3 Patient mortality processes and the Patient Survival probability

Here we present a model for patient survival during (and after) therapy. Patient mortality is modelled as a Poisson process with 3 causes of death: age, treatment related (toxicity) and tumour induced mortality. For simplicity we assume that mortality processes are independent. This framework provides the basis for deriving the CLP which can be generalised to include other events.

We consider a patient with a generic tumour treated with a single chemotherapeutic drug, though the framework is identical for multiple drugs. Patient mortality is described by three independent Poisson processes: tumour-induced mortality with rate *µ*_1_(*N*) that is dependent on the current tumour size, drug-related mortality (i.e. TRM) with rate *µ*_2_(*u*) that is dependent on the current drug concentration, and age-related mortality representing a baseline risk with rate *µ*_*a*_(*t*). Here, *a* denotes the patient’s age at the start of treatment (*t* = 0), so that *µ*_*a*_(*t*) = *µ*_0_(*a* + *t*). The rates *µ*_1_(*N*), *µ*_2_(*u*) and *µ*_*a*_(*t*) are non-negative functions. In the absence of drug or if the tumour is eradicated, the respective mortality rates are equal to zero (*µ*_2_(0) = 0 and *µ*_1_(0) = 0). More complex TRM and tumour-induced mortality models can be used, for instance with a history dependence.

To describe the baseline age-related mortality of an individual, we employ a Gompertz-Makeham (GM) distribution, a distribution that has been used in a number of studies to analyse survival data [9, 18, 36]. The cumulative distribution function (CDF) is,

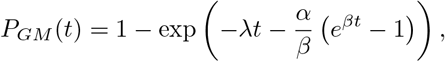

where the parameter *λ* captures baseline (age-independent) mortality, and the parameters *α* and *β* are the scale and shape parameters of age-related mortality, respectively. Provided that an individual is alive at age *a*, the probability of surviving to some future time *a* + *t* is given by

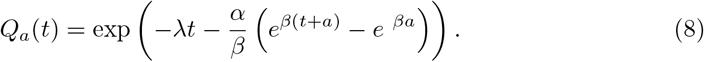

We estimated the parameters *λ, α* and *β* by fitting *Q*_0_(*t*) to national life tables from the office of national statistics [53]; *Q*_0_(*t*) closely reproduces the survival trends in this dataset, Figure 1 **A**. Details on the parameter estimation procedure are provided in Supplement C. Figure 1 **B** and **C** show the expected lifespan and the life expectancy of an average individual in the UK, derived from the distribution *Q*_*a*_(*t*), *a* ∈ [60, 100]. In absence of tumour and drug, the CLP of an individual aged *a* years coincides with the life expectancy shown in Figure 1 **C**.

**Figure 1.**
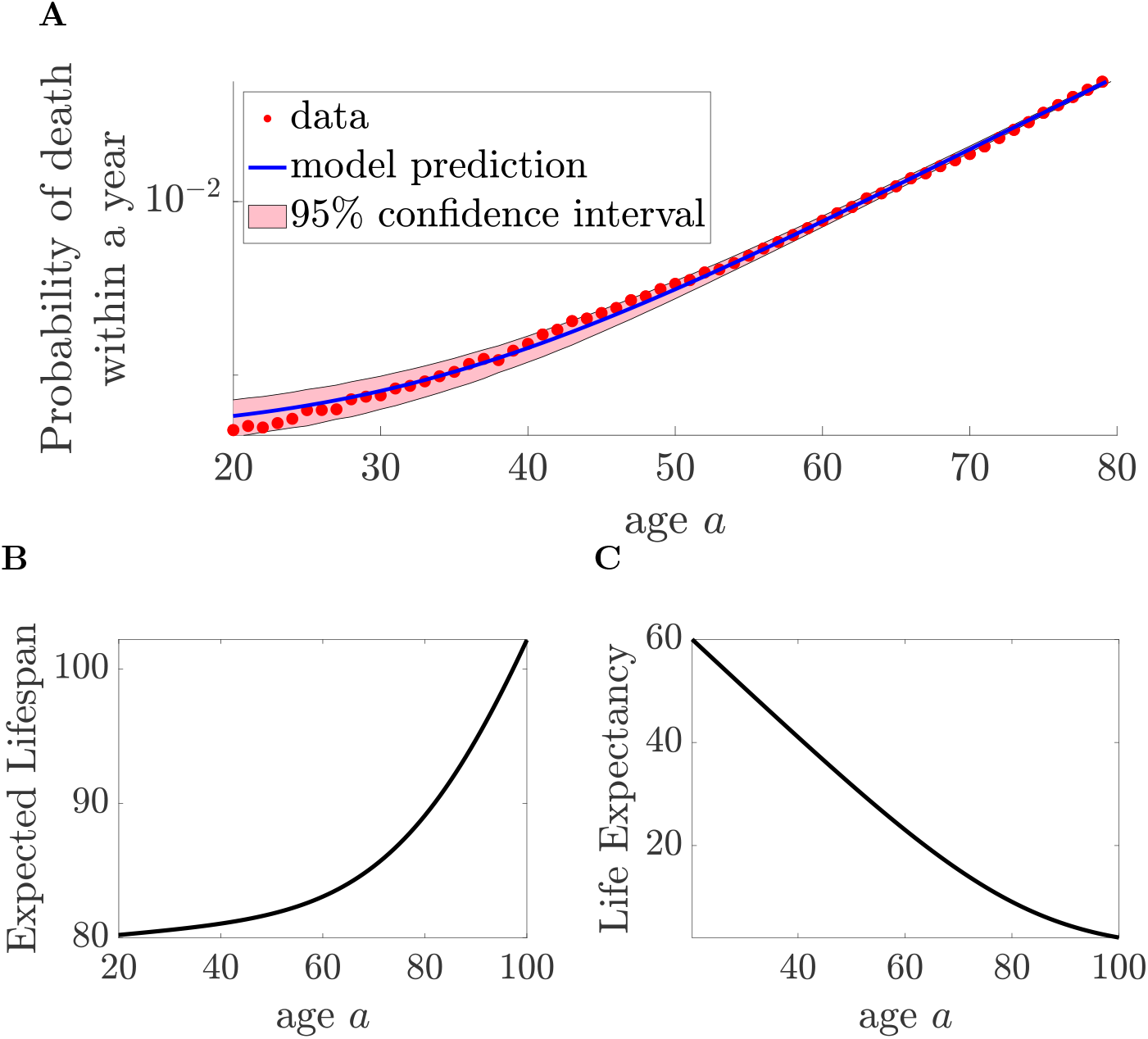
Human age related mortality follows the Gompertz-Makeham distribution. **A.** The probability that an individual of age *a* dies within a year, as a function of age *a*. Red dots represent data, the blue line illustrates the fit and the 95% confidence interval is coloured in pink. Data are taken from [53] and have been smoothed using a three year moving average. **B**. The expected lifespan and **C**. the life expectancy of an average individual (identical to the payoff for tumour free untreated patients CLP[0, 0]) as a function of their age.

The mortality rate at time *t* of a patient aged *a* at the start of therapy (*t* = 0) with current tumour size *N*(*t*), drug dose *u*(*t*) is given by,

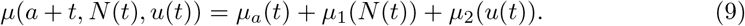

with

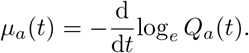

The PDF of the time of death for a patient of age *a* is then given by,

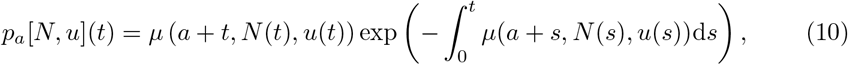

the notation *p*_*a*_[*N, u*](*t*) indicates that the PDF explicitly depends on tumour trajectory *N*(*s*) and treatment history *u*(*s*), with *s* ∈ (0, *t*]. The probability that a patient of age *a* at the start of therapy survives to time *t* is given by

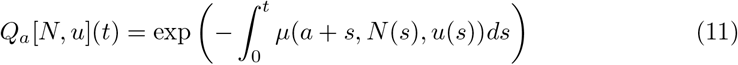

and the corresponding probability of death by time *t*,

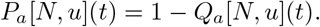

We denote by 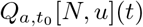 the survival probability of a patient at time *t* who had survived to *t*_0_, with *t*_0_ *< t*. It is defined as

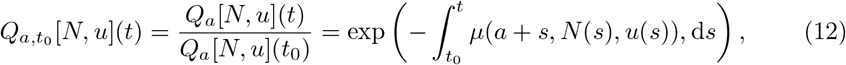

where the expression on the right-hand side follows from conditioning on survival up to time *t*_0_. The following property then holds,

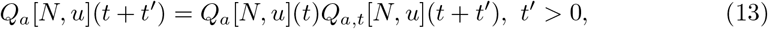

which will be used in the following.

## 4 The Continuous Lifetime Payoff and clinical outcomes

The patient’s expected lifetime can now be constructed based on the distributions of the key events discussed above in Sections 2 and 3 - tumour elimination and patient death. We will be interested in 3 clinical outcomes and their associated probabilities: patient cure (tumour elimination and the patient survives treatment), patient survival but the tumour remains (and thus the patient will relapse) and patient death during treatment. Because of age related mortality and relapse (tumour-induced mortality), the outcome probabilities are dependent on the time when outcome is recorded post-treatment.

We formulate the expected lifetime of a patient undergoing chemotherapy treatment within a fixed time period *I* = [0, *T*_0_], *T*_0_ *>* 0. To simplify notation, we extend the domain of *u*(*t*) defining *u*(*t*) = 0 for all *t > T*_0_. To determine the patient’s expected lifetime, one must average over the two possible outcomes: the tumour being eradicated or its persistence. The probability that the tumour gets eliminated by time *t >* 0 is given by CEP[*N, u*](*t*), Equation 6) and the probability that the tumour eventually gets eliminated is given by the PTE defined as PTE[*N, u*] = lim_*t*→∞_ CEP[*N, u*](*t*), (Equation 7). Recall that PTE[*N, u*] = CEP[*N, u*](*T*_*ζ*_), where *T*_*ζ*_ *> T*_0_ is sufficiently large so that the average tumour size at *T*_*ζ*_ satisfies *N*(*T*_*ζ*_) *> ζK*, for all possible treatments. Let *τ* ∈ (0, ∞) be the tumour elimination time, with conditional PDF (conditioned on elimination),

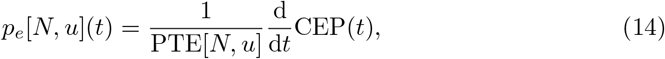

derived in Section 2. If the tumour is eliminated at time *τ*, we set *N*(*t*) = 0 for all *t > τ* in the mortality Poisson process. We define

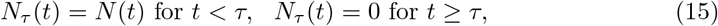

where the trajectory *N*(*t*) has dynamics given in Equation 1.

### 4.1 Survival Time under Tumour Elimination and Persistence

In the case where the tumour does not get eliminated, the expected lifetime is given by:

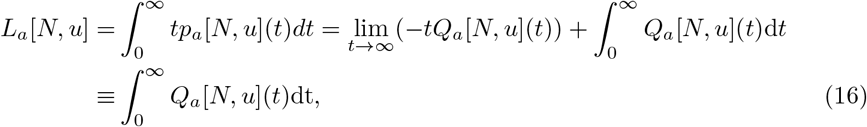

where *p*_*a*_ (Equation 10) and *Q*_*a*_[*N, u*](*t*) (Equation 11) are respectively the PDF of the mortality Poisson process and the survival probability for a patient of age *a* undergoing chemotherapy treatment. *L*_*a*_[*N, u*] is finite since the death rate is bounded from below as follows from the Gompertz-Makeham distribution, *i*.*e*. patients always die (thus *Q*_*a*_[*N, u*](*t*) *< e*^−*λt*^ → 0 as *t* → ∞). This expression for *L*_*a*_[*N, u*] is an approximation since *N*(*t*) represents the average tumour size, averaged over all trajectories including those where the tumour is eliminated. However, since trajectories in which the tumour is eventually eliminated contribute negligibly to the average tumour size *N*(*t*), *L*_*a*_[*N, u*] still provides a sufficiently accurate estimate of the expected lifetime under the condition of no tumour elimination. Using Equation 13, *L*_*a*_[*N, u*] can also be expressed as:

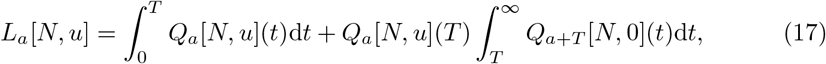

For the case where the tumour gets eliminated at time *τ >* 0 (finite), the expected lifetime is given by

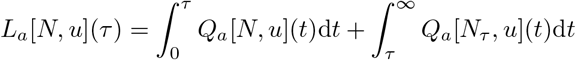

where *N*_*τ*_ is defined in Equation 15, the tumour trajectory when the tumour gets eliminated at time *τ*. Since the mortality rate is additive across the 3 mortality processes (independence), we have for *t > τ*,

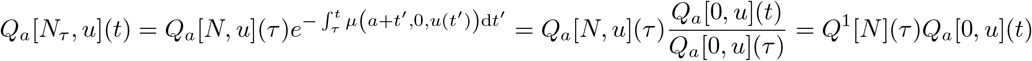

where

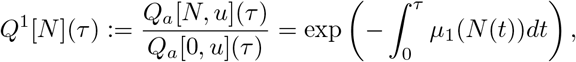

represents the survival probability of tumour-induced mortality only. This is a Poisson process with rate *µ*_1_(*N*), and PDF *p*^1^[*N*](*t*) = *µ*_1_(*N*(*t*))*Q*^1^[*N*](*t*). We then obtain the expression:

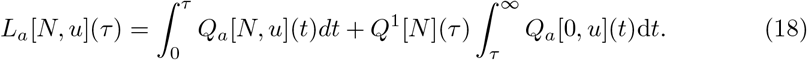

Since 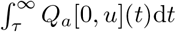 has an upper bound and lim_*τ*→∞_ *Q*^1^[*N*](*τ*) = 0, it follows that

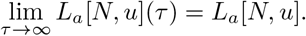

Thus, in terms of survival time, tumour persistence can be seen as the limiting case of tumour elimination with elimination time *τ* → ∞.

### 4.2 Derivation of the CLP

Averaging over tumour persistence (with probability 1 − PTE[*N, u*]) and tumour elimination time (with elimination time PDF PTE[*N, u*] *p*_*e*_[*N*_*u*_](*t*)), the expected lifetime is given by:

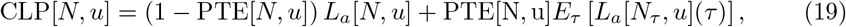

where the expectation is taken over the distribution of the elimination time *τ* ∈ [0, ∞) conditioned on elimination, *i*.*e*. the PDF *p*_*e*_[*N, u*](*t*) (Equation 14),

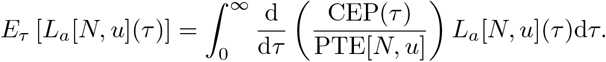

For any *T ≥ T*_0_, the CLP can be expressed explicitly in terms of the CEP, the survival probabilities and their derivatives,

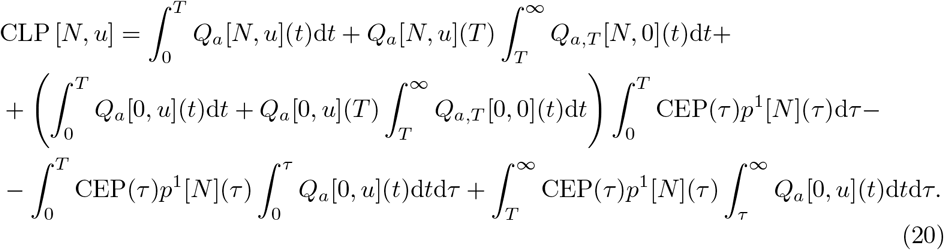

Details of the derivation can be found in Supplement D.

To use the CLP as a payoff function of a finite-horizon optimal control problem, it is necessary to derive a closed-form expression for the integrals that extend to infinity. It is important to note, however, that some of these integrals in Equation 20 depend explicitly on the tumour size *N*(*t*), that itself depends on the history of the drug. By setting *T* = *T*_*ζ*_ (with *N*(*T*_*ζ*_) = *ζK* and *ζ*1) the tumour size *N*(*t*) may be approximated by the (constant) carrying capacity *K* for *t* ≥ *T*_*ζ*_, allowing the integrals to be evaluated in closed form:

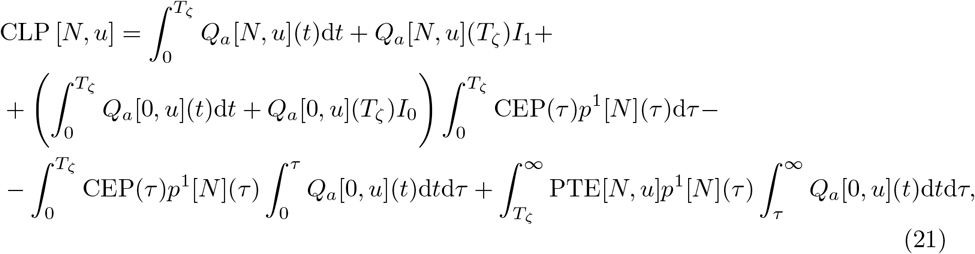

where 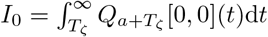 and 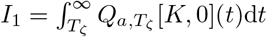 are positive constants for given values of *T*_*ζ*_, and can be expressed in the closed forms,

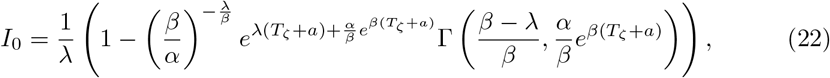

and

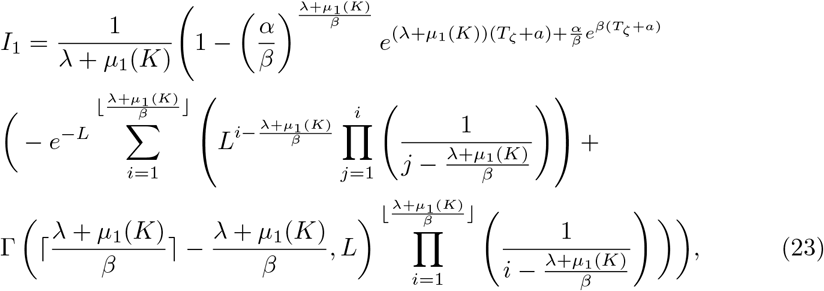

with 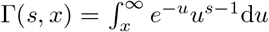 the incomplete lower gamma function and ⌊*x*⌋, ⌈*x*⌉ the lower and upper integer parts of *x* ∈ *R*. The derivation of Equations 22 and 23 can be found in Supplement E. Then by rearranging terms (details in Supplement F), the CLP can be expressed in the Mayer form:

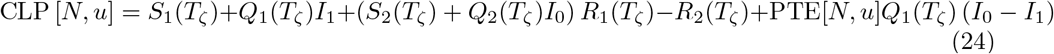

where

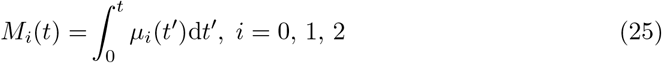

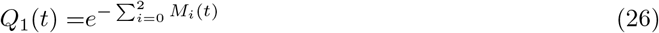

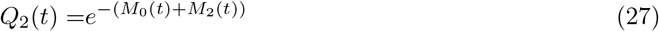

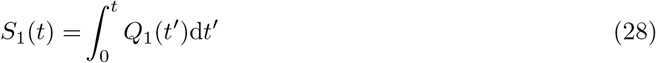

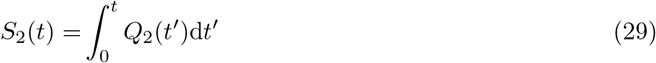

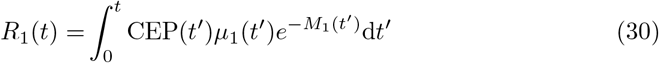

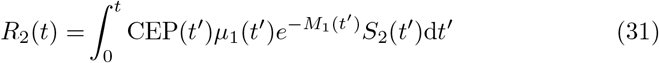

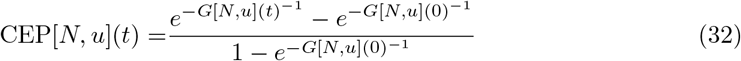

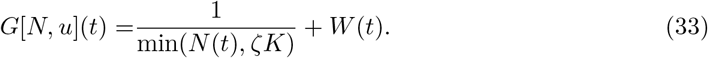

We emphasize that, under the requirement *N*(*T*) ≥ *ζK*, the CEP remains invariant for *t* ≥ *T*, yielding PTE[*N, u*] = CEP[*N, u*](*T*_*ζ*_).

Here we have separated the two roles of the horizon time in a classic control theory problem, specifically the time interval over which the control is to be determined and the interval over which the objective is defined. In the following we refer to *T*_0_ as the control horizon, the predetermined interval [0, *T*_0_] where the control is to be determined, and *T*_*ζ*_ as the objective horizon. Recall that the interval [*T*_0_, *T*_*ζ*_] serves to ensure that the condition *N*(*T*_*ζ*_) ≥ *ζK* is satisfied for every possible treatment trajectory *u*(*t*). The parameter *T*_*ζ*_ can thus be chosen arbitrarily large without increasing the computational complexity of the optimisation. The CLP only depends on *T*_0_ through the constraint on the control.

Our finite time horizon control problem can now be stated as the following:

**Find the control** *u*(*t*) ∈ [0, 1] **in interval** *t* ∈ [0, *T*_0_] **that maximises the CLP, Equation 24**, **subject to the dynamics, Equation 1**, **5**, **and determine the probabilities of the 3 outcomes: cure, relapse and death due to drug-related mortality**.

### 4.3 Clinical outcomes

There are three endpoints of interest, Table 1, patient cure (survival and elimination), TRM and surviving treatment but later the tumour relapses. We define cure and TRM events conditional on not dying of age-related mortality; they are then independent of any specific time after treatment completion. We define the completion time of therapy,

**Table 1.** Probability of each clinical endpoint: (i) cure, conditional on not dying of age-related mortality, defined as the probability the tumour is eliminated and the patient does not die of drug-related mortality; (ii) drug-related mortality (TRM); and (iii) the patient survives to the completion of treatment (*T*_*complete*_) but succumbs to tumour-induced mortality (relapse), the product of not eliminating the tumour and surviving to *T*_*complete*_ (conditioned on no elimination). Conditional cure and drug-related mortality are in fact independent of *T*_0_; any time ≥ *T*_*complete*_ can be used since *u* = 0 for *t > T*_*complete*_.

| Clinical endpoints | Probability |
| --- | --- |
| Cure (conditional) | $\frac{Q_a[0, u](T_0)}{Q_a[0, 0](T_0)} \int_0^\infty Q^1[N](\tau) \left( \frac{dCEP(\tau)}{d\tau} \right) d\tau$ |
| Drug-related mortality | $1 - \frac{Q_a[0, u](T_0)}{Q_a[0, 0](T_0)}$ |
| Survived treatment, dies from tumour | $(1 - PTE[N, u]) Q_a[N, u](T_{complete})$ |

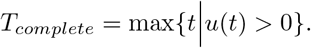

Since mortality processes are independent, we can compute the associated clinical outcomes at any specified time by multiplying by the age-related mortality survival probability (if TRM can occur post *T*_*complete*_ these conditional survival at a specified time need modifying). So the probability of cure, surviving to *T*_*complete*_ and the tumour being eliminated (possibly beyond *T*_*complete*_) is,

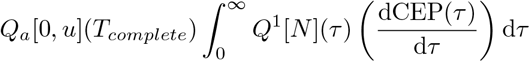

where we used the fact that drug-related mortality can only occur in [0, *T*_*complete*_], so are free to choose the time when we define the cure conditional on not dying of age in Table 1. Finally we define the probability that a patient who survived treatment will succumb to tumour-induced mortality in absence of any further treatment.

## 5 Case study with logistic tumour growth

We demonstrate the CLP framework determining the optimal chemotherapy strategies for a tumour growing with logistic growth dynamics with three possible outcomes: cure, TRM and failure to clear the tumour.

The CLP (Equations 24-33) is maximized assuming logistic tumour growth under the influence of a cytotoxic drug. Tumour dynamics is:

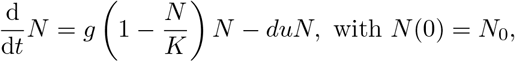

with *g* the maximum growth rate of cancer cells and *K* the carrying capacity, both in the absence of drug. For simplicity we scale the control *u*(*t*) to 1 (*i*.*e*. 0 ≤ *u*(*t*) ≤ 1, *∀t* ∈ [0, *T*]) so that *u* = 1 is the MTD. To define the TCP we need to specify an underlying branching process - we assume the birth rate is *a*(*t*) = *g* and death rate 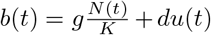, where the first term captures death due to intercellular competition and the second term drug induced death. The tumour growth parameters are taken from [5], with *g* = 0.502 days^−1^ and *K* = 1.297 × 10^9^ cells, whereas the per capita death rate of cancer cells due to the drug is set to *d* = 1 day^−1^, satisfying *d > g* for an effective drug (at MTD, the tumour size *N* → 0 as *t* → ∞).

We will use linear approximations for the mortality rates for simplicity. The tumour-induced patient mortality rate is linear in the tumour size,

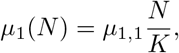

scaled by the carrying capacity. The tumour-induced mortality rate of a tumour at carrying capacity (*N* = *K*) is set to 1*/*60 days^−1^, corresponding to a mean survival time of approximately 2 months in line with reported survival times in terminal cases [3, 35]. The treatment related mortality rate is assumed proportional to the drug concentration,

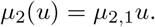

The parameter *µ*_2,1_, hereafter referred to as the *drug sensitivity coefficient*, captures patient susceptibility to treatment related toxicity.

Details of the numerical method and its implementation to solve the optimal control problem stated in Section 4(b), are provided in Supplement G. In brief, the payoff is maximized using a standard direct method [46], in which the control function is approximated by piecewise-constant functions. This transforms the optimal control problem into a nonlinear programming (NLP) problem, where the decision variables (with respect to which the objective function is maximised) are the discretisation time points (the time instances where the control is allowed to change) and the corresponding control values between these points. Here, we discretise the control using 30 discretisation points with a (control) time horizon *T*_0_ of 60 days.

All numerical solutions of the optimal control problem were found to be bang-bang with at most one switch (from MTD to no drug). Specifically we observed 3 types of optimal solutions: no treatment, MTD throughout interval [0, *T*_0_] and treat-and-stop (drug administration at MTD over [0, *x*] with *x < T*_0_). How these optimal solutions emerge as a balance between a reduction in the tumour induced mortality and the probability of surviving treatment can be understood by examining the CLP under variation of MTD drug administration period *x* with an increasing level of drug tolerance, Figure 2. Drug tolerance is quantified by *µ*_2,1_. When drug tolerance is poor, (*µ*_2,1_ large, Figures 2 **A** and **B**), treating the patient for any period *x >* 0 does not extend the life expectancy since the drug induced mortality is too high. No treatment is thus the optimal strategy. At a critical value of *µ*_2,1_, MTD treatment for a specific time period emerges as a local optimum of the treated solutions that yields the same payoff as the no-treatment solution, (Figure 2 **C**). Drug induced mortality is thus balanced by the increased life expectancy of those that survive. All other MTD-based strategies remain suboptimal (Figure 2 **C**). As drug tolerance increases (decreasing *µ*_2,1_ below this critical threshold), strategies with MTD for a specific (optimal) time period become optimal (Figure 2 **D**). For this case, if the horizon time *T*_0_ is below the optimal drug administration period the optimal strategy is either no treatment or MTD for the full horizon.

**Figure 2.**
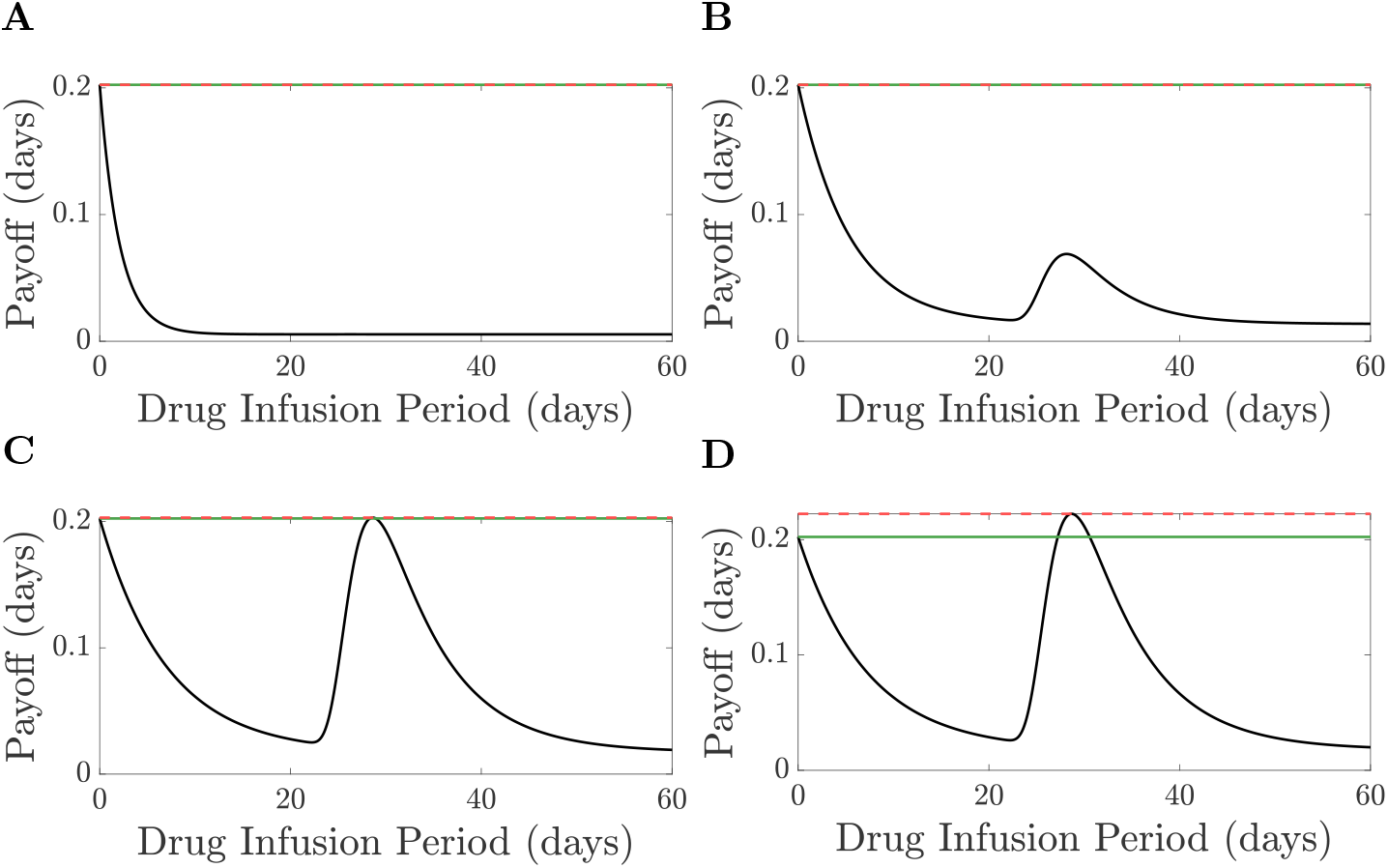
Payoff as a function of the drug infusion period *x*. **A.** *µ*_2,1_ = 1*/*2 days^−1^, **B**. *µ*_2,1_ = 1*/*5 days^−1^, **C**. *µ*_2,1_ = 1*/*6.36 days^−1^, **D**. *µ*_2,1_ = 1*/*6.5 days^−1^. Continuous black lines show MTD drug regiment for administration period *x*, the red dashed line indicates the maximum payoff whereas the green continuous line indicates the value of the payoff when no drug is administered. The initial tumour size is *N*_0_ = 10^6^ cells, the carrying capacity is *K* = 1.297 10^9^ cells and *µ*_1,1_ = 1*/*60 days^−1^. The per capita tumour growth in absence of the drug is *g* = 0.502 cells/days and the drug induced death rate of cancer cells is *d* = 1 day^−1^.

Treat-and-stop solutions involve a drug administration period *x* long enough to substantially increase the probability of tumour elimination, extending the lifetime of cured patients thereby offsetting the risk of treatment-related mortality. This is illustrated in Figure 3 where we analyse the impact of MTD duration (*x*) on the PTE, the probability of patient survival and the patient’s expected lifetime. Crucially, only when the PTE rises does a patient’s expected lifetime increase sharply with the drug administration period Figure 3 **A**. Extending MTD administration beyond the optimal 32 day period yields little additional improvement in PTE whilst it impacts treatment related mortality, the survival probability *Q*_*a*_[*N, u*](*x*) decreasing. Beyond 70 days MTD, the survival of (treated) patients without a tumour (*Q*_*a*_[0, *u*](*x*)) approaches the overall survival of patients with a tumour, Figure 3 **B**, *i*.*e*. tumour-induced and age-related mortality are negligible. Consequently, such prolonged regimens result in suboptimal expected lifetimes, Figure 3 **A**.

**Figure 3.**
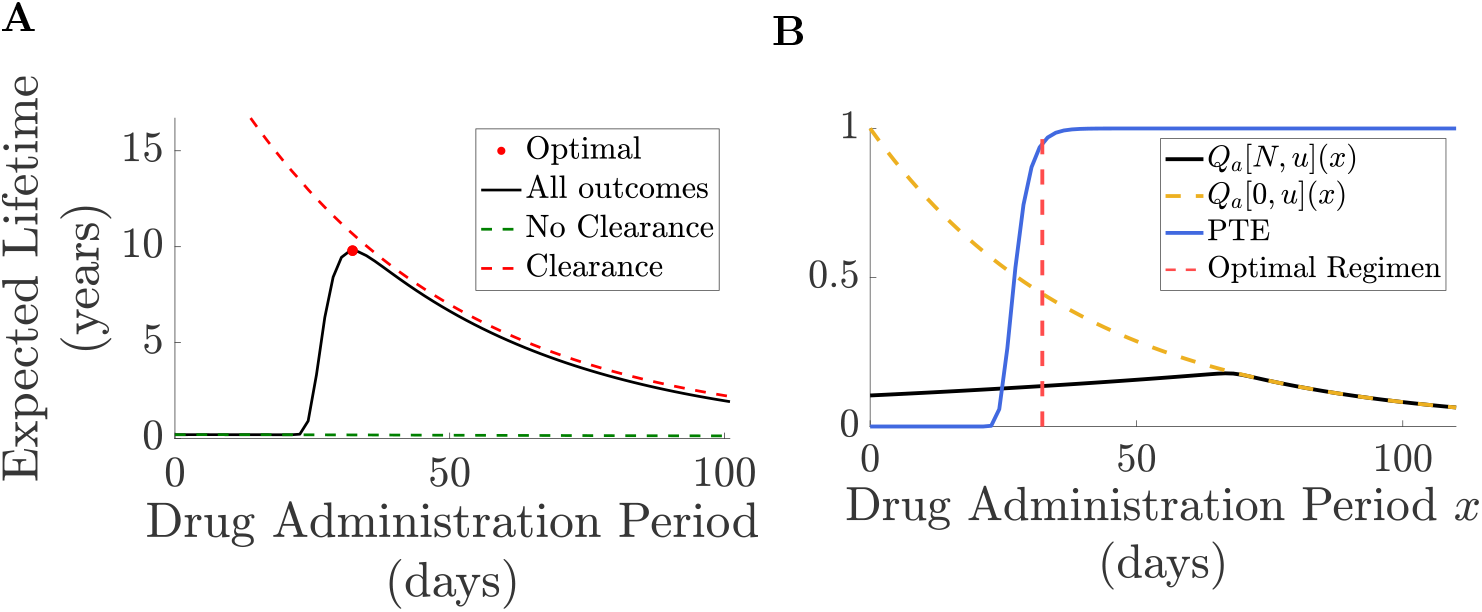
Survival curves with MTD duration. **A:** Life expectancy under continuous MTD versus drug administration period. The red dot indicates the optimum; green and red dashed lines show life expectancy conditional on treatment failure and success, respectively and the black line represents the CLP (expected lifetime averaged over all outcomes). **B:** Survival probabilities and PTE under continuous MTD treatment as a function of the drug administration period *x*. The blue line shows the PTE, while the yellow dashed line and black line represent, respectively, the probability of surviving drug administration beyond time *x* (*Q*_*a*_[0, *u*](*x*)), and the probability of surviving beyond time *x* given that the tumour persists at the end of the treatment (*Q*_*a*_[*N, u*](*x*)). The red vertical line indicates the optimal drug administration regimen. Patients are 60 years old with an initial tumour size of 10^6^ cells. Parameters are as in Figure 2 with 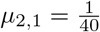. *T, T*_0_ are 600 and 110 respectively.

The survival curves under the optimal drug regimen is shown in Figure 4. We separate causes of patient death (age-related, tumour-induced, or drug-related) comparing the survival probability *Q*_*a*_[*N, u*](*t*) across three idealised scenarios: (i) an individual with no initial tumour and no treatment (*Q*_*a*_[0, 0]); (ii) an individual with no tumour receiving optimal treatment for a tumour sized *N*_0_, (*Q*_*a*_[0, *u*]); and (iii) an individual with an initial tumour of size *N*_0_ receiving optimal treatment (*Q*_*a*_[*N, u*]). These are all shown in Figure 4. Recall that once the tumour is eliminated the tumour size is set to zero. Consequently *Q*_*a*_[0, *u*] represents the survival probability of a patient whose tumour was eliminated at time *t* = 0, and is therefore an upper bound for the survival probability of cured patients (with initial tumour size *N*_0_). Figures 4 **A** and **B** depict survival probabilities for 60-year-old and 95-year-old individuals, respectively. During treatment drug-related mortality dominates survival, with the survival probability of an uncured individual (*Q*_*a*_[*N, u*](*t*)) being indistinguishable from the upper bound of a cured individual (*Q*_*a*_[0, *u*](*t*), for *t* ∈ [0, *x*]) whilst age related mortality is negligible in that interval. Notably, there is no significant increase in CEP during the first 20 days of therapy because the tumour remains too large (the tumour extinction rate is small); however, in the final 10 days, the CEP rises sharply, eventually exceeding 0.95 (for these parameters). After therapy the survival probability of cured patients declines slowly (being identical to *Q*_*a*_[0, *u*](*t*) since the tumour was eliminated) because of age-related mortality. In contrast, nearly all patients die within 9 months post-treatment if the tumour was not eradicated (*Q*[*N, u*](*t*)), with a survival curve that is delayed relative to no treatment (*Q*[*N*, 0](*t*)). For around a month after treatment ends the survival probabilities for both these groups (cured or retaining a tumour) are nearly identical. This is because there is a delay before the tumour regrows to a size that significantly impacts survival. This delay is the time taken for relapse to occur: immediately following the drug administration period the tumour burden *N* is small but in the absence of eradication the tumour regrows over time, eventually resulting in patient death (if left untreated). This observation distinguishes the CEP from traditional optimal control formulations. By explicitly accounting for tumour elimination, the CLP provides a more informative (and clinically relevant) measure of anti-tumour efficacy than standard metrics, such as tumour size at the end of treatment. On these parameter values age of the patient had little impact with the optimal treatment strategy and survival dynamics being similar for individuals aged 60 and 90 years. The main difference is that, post-treatment, survival steadily declines for a cured 90-year-old due to age-related mortality, while it plateaus for a cured 60-year-old, Figure 4.

**Figure 4.**
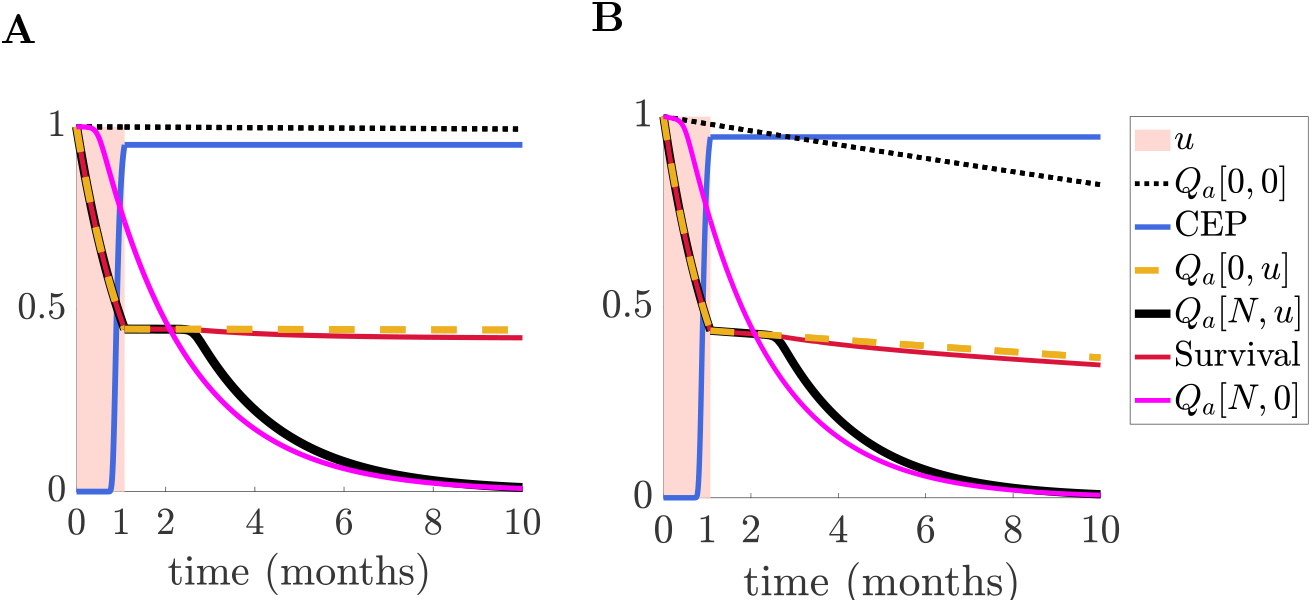
Decomposition of the survival curve under the optimal drug regimen as a function of time. **A.** A 60 year old individual and **B**. A 95 year old individual. The area coloured in pink illustrates the time period when MTD is administered. The dotted black line illustrates the survival probability of an average individual in the population without a tumour or treatment (*Q*_*a*_[0, 0]), the dashed yellow line illustrates the probability of an individual (with no tumour) surviving drug administration (*Q*_*a*_[0, *u*]) whereas the continuous black line illustrates the survival probability of an individual in the case were the tumour does not get eliminated (*Q*_*a*_[*N, u*]). The blue continuous line is the CEP, the red line the overall survival under optimal treatment and the magenta line shows the survival probability in the complete absence of treatment. Parameters are as in Figure 3.

The CEP and the survival probabilities together determine patient overall survival (OS) under treatment, a standard measure used to assess (and compare) therapy. OS at time *t* ∈ [0.∞) is computed as,

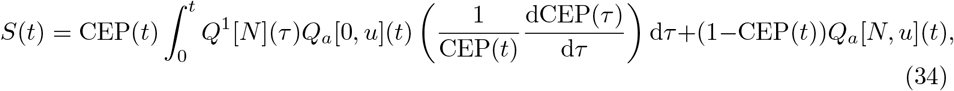

decomposing survival into two components. The first term represents the fraction of patients who have been cured by time *t*: it combines the CEP with the survival probability averaged over all possible elimination times. The second term is the expected fraction of patients who are still alive at time *t* but are not cured (referred to as uncured). In absence of drug administration the CEP is approximately zero (spontaneous cure is negligible) and overall survival is well approximated by *Q*_*a*_[*N*, 0]. Figure 5 shows the OS curves for patients receiving continuous MTD for 28 days and under the optimal treatment strategy (32 days MTD), decomposed into cured patients and those that retain the tumour and survive to time *t*. There is a sharp decline in OS during treatment because of treatment related mortality, Figure 5. For most of the treatment period, the majority of patients remain uncured. However, near the end of therapy, the CEP increases steeply, leading to a rapid rise in the treated (cured) fraction and a corresponding decline in the uncured fraction. After treatment ends, the cured fraction remains essentially constant, as the CEP changes very little and the probability of post-treatment cure is negligible. The uncured (surviving) fraction initially stabilizes, Figure 5 **A**, but tumour relapse progressively reduces survival in this group. Under the 28 day MTD administration period (Figure 5 **A**), nearly all untreated patients die by nine months, and OS approaches the fraction of cured patients (approximately 32%). Under the optimal strategy (Figure 5**B**), with 42% cured patients, the vast majority of patients that survive treatment are cured, so the post-treatment decline in OS is much less pronounced than under 28-day continuous MTD (Figure 5**A**).

**Figure 5.**
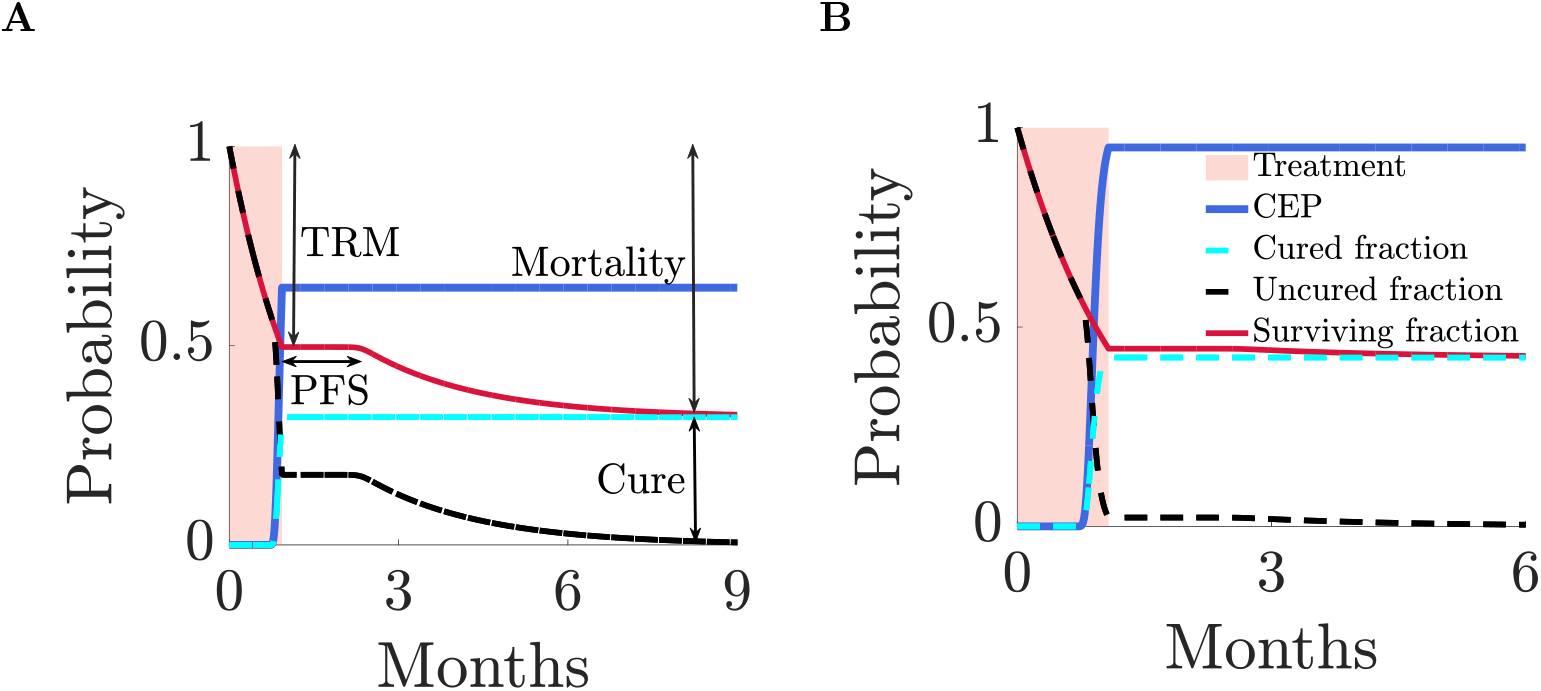
Survival and outcome for a 60-year-old patient undergoing therapy. **(A)** Continuous MTD for 28 days, highlighting treatment-related mortality (TRM), progression-free survival (PFS, the period before disease progression or death), the cure fraction and the mortality fraction (the proportion that die in the ‘long-term’ relative to disease dynamic time scales). **(B)** optimal treatment (32 days MTD). The shaded region denotes the treatment period, while the unshaded region corresponds to follow-up. The solid blue curve represents the tumour elimination probability (CEP). The cyan dotted curve shows the proportion of patients with the tumour eliminated, and the dark dotted curve shows the proportion with the tumour persisting. The solid red curve represents overall survival. The tumour will be undetectable for some of that period. Parameter values are as Figure 3.

Age has a limited effect on the structure of the optimal drug regimen which primarily depends on the initial tumour size and the drug sensitivity coefficient *µ*_2,1_. For example, the optimal drug administration period for a 90-year-old individual, with various initial tumour sizes and a drug sensitivity coefficient of *µ*_2,1_ = 1*/*40 days^−1^, almost overlaps with that of a 20-year-old individual with the same tumour burden and drug sensitivity, Fig. 6 **A**. The relative survival (the life expectancy of a treated individual relative to the life expectancy of an average healthy individual of the same age) are also nearly identical, Fig. 6 **B**. In contrast, a 90-year-old patient with a lower drug sensitivity coefficient of *µ*_2,1_ = 1*/*80 days^−1^ increases the drug administration period compared to patients with *µ*_2,1_ = 1*/*40 days^−1^, which results in a markedly higher life expectancy than patients with a higher drug sensitivity coefficient.

**Figure 6.**
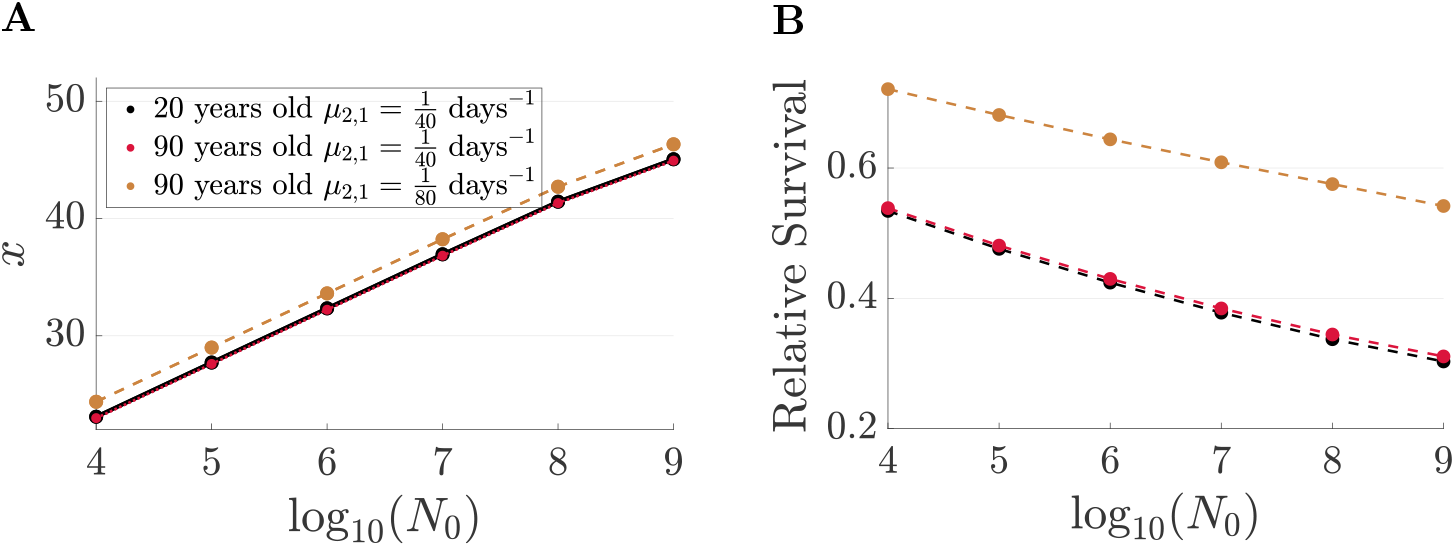
Impact of tumour size *N*_0_ on the optimal drug administration period and the relative life expectancy. **A** the optimal drug administration period *x* as a function of log_10_(*N*_0_), where *N*_0_ is the initial tumour size and **B** the life expectancy (averaged over all outcomes) of an individual after optimal treatment, relative to the life expectancy of an average individual as a function of log_10_(*N*_0_). Black colours relate to 20 year old individuals with drug induced death rate per day of MTD administration equal to *µ*_2,1_ = 1*/*40, red colours relate 90 year old individuals with *µ*_2,1_ = 1*/*40 and brown colours correspond to 90 year old individuals with *µ*_2,1_ = 1*/*80.Remaining parameters are as in Figure 2.

In the following analysis, the patient’s age is fixed at 60 years, while the initial tumour size (*N*_0_) and the drug sensitivity coefficient are varied. We quantify the patient’s drug tolerability by the inverse of the drug sensitivity coefficient, 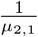. This is the expected survival time of a patient on MTD conditioned on not succumbing to other causes of mortality. To efficiently explore a wide range of *N*_0_ and drug tolerances, we use only two discretisation time points to optimise the CLP in the numerics adequate to model treat-and-stop solutions. However, as shown in Supplement H, the optimal solutions closely match those obtained using 30 discretisation time points.

The optimal drug administration period (*x*_opt_) depends primarily on the initial tumour size (*N*_0_), with a weaker but still positive dependence on the patient’s tolerance to the drug (1*/µ*_2,1_) Figure 7**A**. It increases monotonically with both quantities. The upward trend of *x*_opt_ with increasing patient drug tolerance is intuitive: patients who are less susceptible to drug toxicity can safely receive treatment over longer durations. The influence of *N*_0_ on *x*_opt_ arises from the fact that larger tumours require a longer drug administration duration to eliminate the tumour and achieve a reasonable PTE.

**Figure 7.**
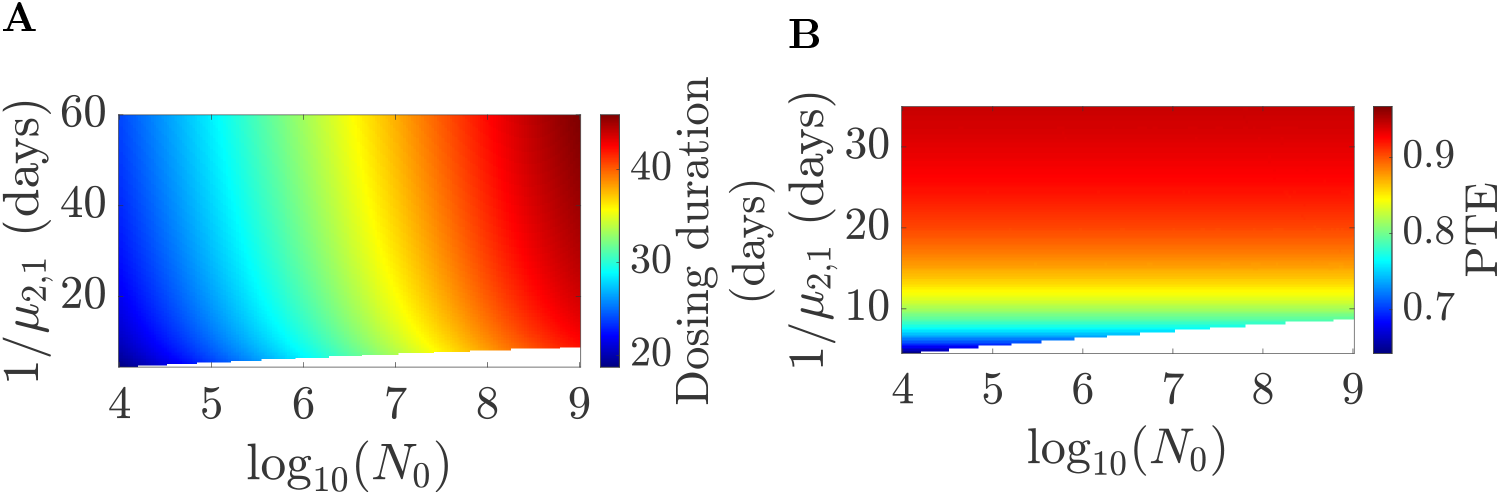
Optimal drug administration period and PTE with drug tolerance and *N*_0_ **A** the optimal drug administration period and **B** the PTE of 60 year old patients, as functions of log_10_(*N*_0_) (*N*_0_ the initial tumour size) and the drug tolerance interval 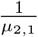. Remaining parameters are as in Figure 2.

The optimal PTE is set by the patient’s tolerance to the drug, increasing with 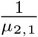, Figure 7**B**, being practically independent of *N*_0_. PTE values range from 0.64 to 0.96 with the lower PTE values corresponding to treatment regimens curtailed by rising drug toxicity.

Based on the tumour burden at detection and on the drug tolerability, patients are categorised as either treatable or untreatable (Figure 8). For instance, if 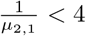 days for an initial tumour size of 10^4^ cells no treatment is optimal, reflecting the inability of the patient to benefit from therapy (Figure 8**A**). The threshold level of the patient’s drug tolerability for treatment increases with tumour size. Crossing this threshold leads to a jump in the optimal drug administration period, jumping from 0 to over 20 days of MTD. However, the expected lifetime under optimal treatment remains continuous across this boundary (Figure 9**A**); continuity is expected because a local maximum emerges in the payoff with MTD duration as seen previously, Figure 2. For patients near the threshold the benefit of treatment is marginal — there is only a slight increase in expected lifetime compared to no treatment.

**Figure 8.**
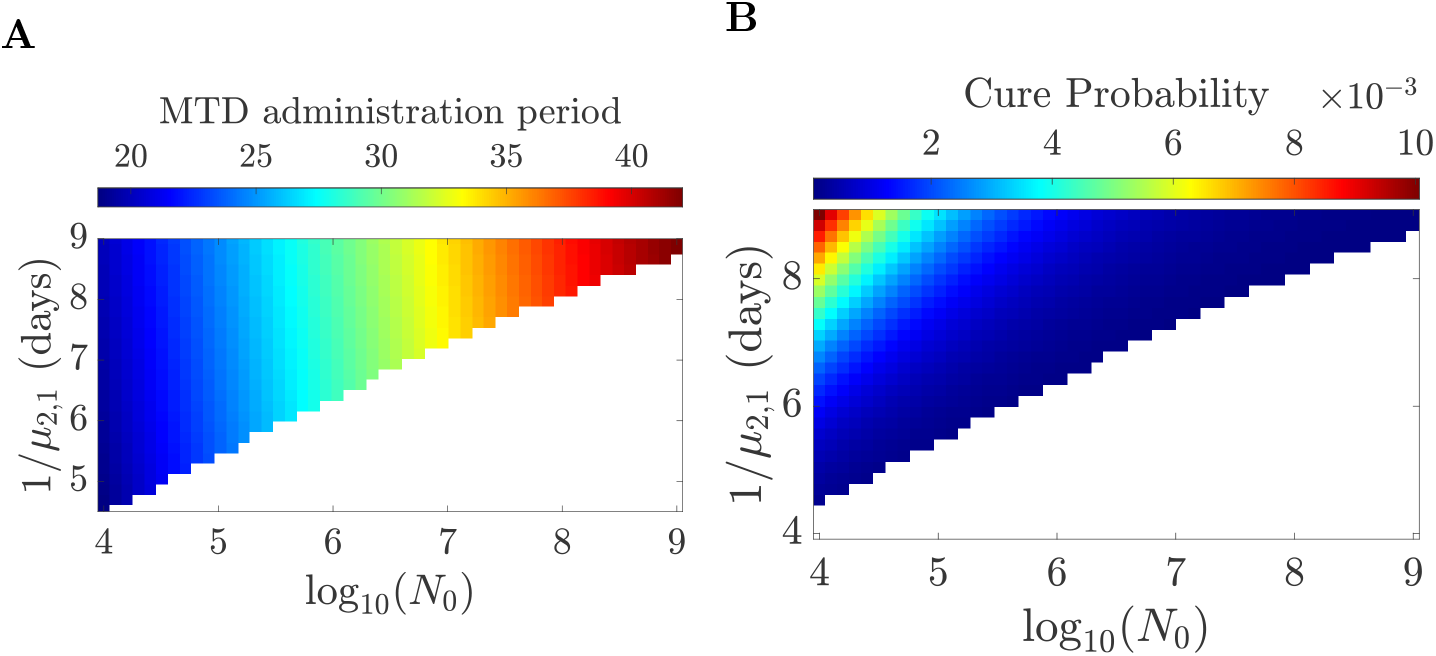
Detail for the optimal drug administration period and the cure probability for 60-year-old patients, as a function of initial tumour size and drug tolerability interval. **A** The optimal drug administration period (detail of Figure 7A) and **B** the cure probability (conditional on not dying of age-related mortality) for 60-year-old patients, as a function of initial tumour size (log_10_-scaled) and drug tolerability interval (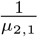). Coloured regions represent patients who can benefit from treatment, while white regions correspond to untreatable cases. Remaining parameters are as in Figure 2.

**Figure 9.**
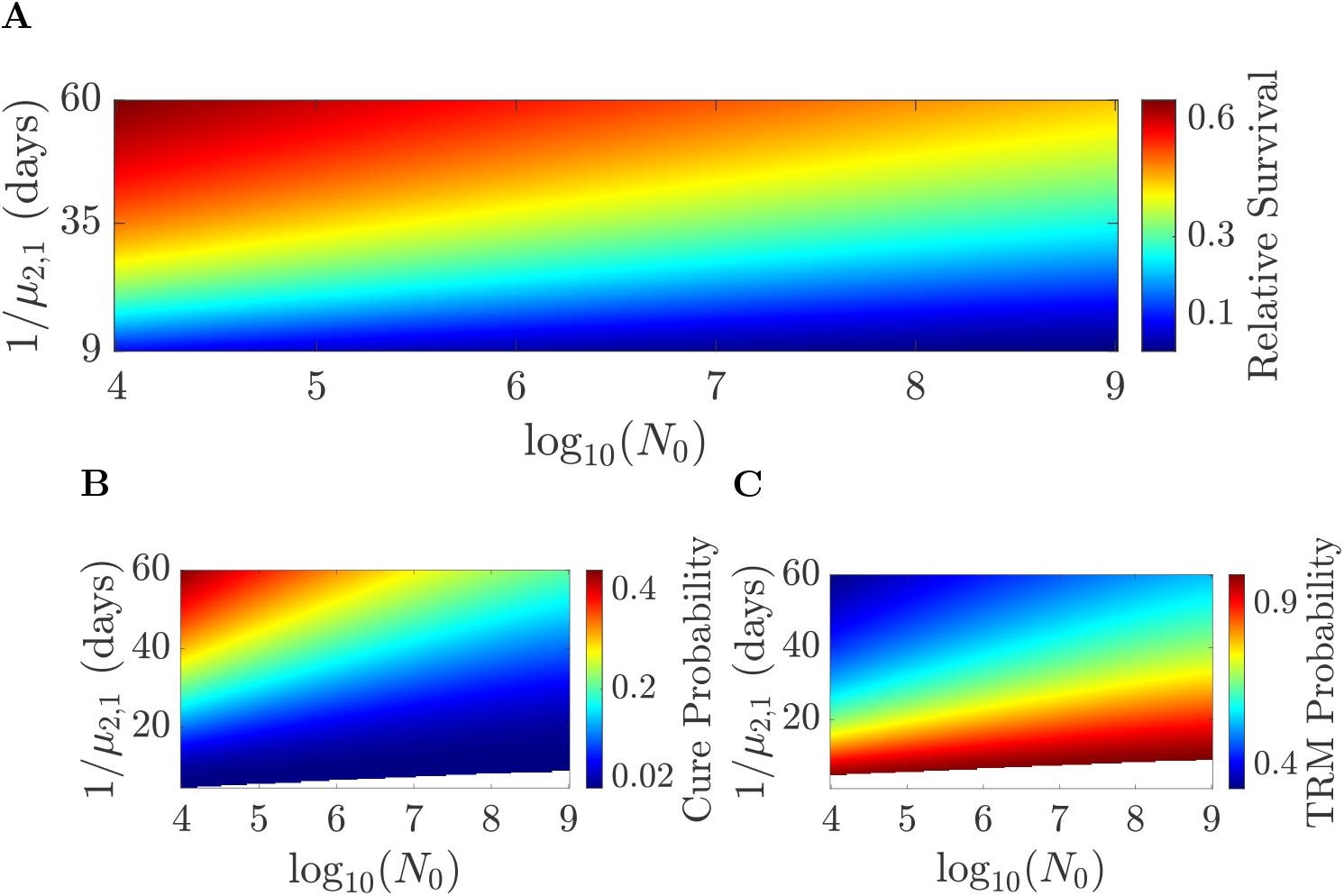
Survival and outcome of a 60-year-old patient under optimal treatment. **A.** The relative life expectancy of a treated 60 year old patient, under optimal treatment, relative to the life expectancy of a healthy 60 year old individual, **B**. the cure probability (conditional on not dying of age-related mortality) and **C**. the drug-related death probability (TRM), of 60 year old patients, as functions of log_10_(*N*_0_) (*N*_0_ the initial tumour size) and the drug tolerability 1*/µ*_2,1_. Parameters are as in Figure 7.

Among treatable patients (for parameters as stated) those with the lowest drug tolerance exhibit a PTE of around 0.64 (Figure 7**B**), a cure probability smaller than 0.01 (Figure 8**B** and 9**B**), and a probability of drug-related mortality exceeding 0.9 (Figure 9**C**). This indicates that optimal strategies favour tumour elimination, even at the cost of a high risk of TRM. Indeed, in the absence of treatment, life expectancy is typically less than three months (Figure 10). In contrast, successful tumour elimination leads to a significant survival benefit: life expectancy (averaged over all outcomes) exceeds 10% of that of an average 60-year-old and may reach over 60% in some cases (Figure 9**A**)

**Figure 10.**
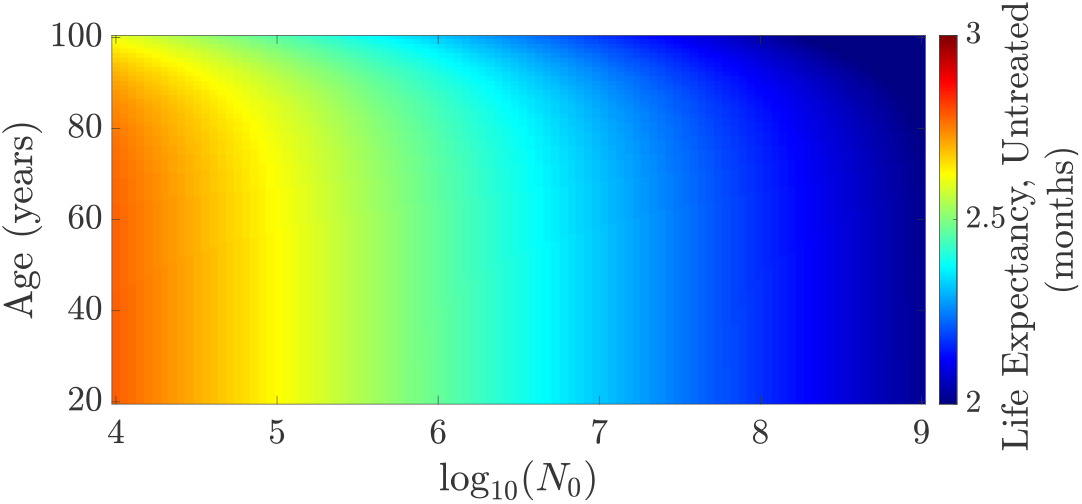
Life expectancy (in months) of an untreated individual as a function of age and initial tumour size (loged in base 10). Parameters are as in Figure 2.

## 6 Parameter estimation from simulated survival curves

All parameters of the CLP have a direct biological interpretation and thus can in principle be estimated from clinical data, for instance from survival curves. Here, we demonstrate likelihood based parameter estimation from OS curves using simulated datasets. Since patients are heterogeneous in practice, i.e. the model parameters will vary across individuals introducing parameter variation in the patient cohort, we demonstrate that optimal solutions are robust to low levels of heterogeneity but performance degrades with the level of heterogeneity as expected. Utilising additional data sources for the inference of tumour dynamics and drug efficacy is indicated.

We analyse inference of 4 model parameters from overall survival (OS) curves: specifically the tumour growth rate *g*, the drug-induced tumour cell death rate *d*, the tumour-related mortality rate coefficient *µ*_1,1_ and the drug-related mortality rate coefficient *µ*_2,1_. There are 3 other parameters we assume known. The baseline mortality rate *µ*_*a*_(*t*) has already been calibrated using life table data, Supplement C and the carrying capacity *K* of the tumour is assumed sufficiently large that it has little impact on the dynamics, *i*.*e*. we assume the initial tumour size is 10^6^ cells, two orders of magnitude below the tumour capacity *K*. Finally, the initial tumour size is assumed to be known by direct measurement, for instance from a computed tomography (CT) scan as appropriate for solid tumours. We simulate OS curves for 500 patients treated with 20 days of continuous MTD using Monte Carlo simulations. In brief, we consider a time horizon of one year and group deaths into daily intervals (*t*_*i*−1_, *t*_*i*_], *i* = 0, …, *k* = 364, *t*_0_ = 0. The probability that a patient dies on day *i* (within interval (*t*_*i*_, *t*_*i*+1_]) is given by *D*_*i*+1_ = *S*(*t*_*i*_) − *S*(*t*_*i*+1_), where *S*(*t*) is given by Equation 34. We used a maximum likelihood estimation methodology, Supplement I, fitting the model’s survival curve *S*(*t*) to the data. We first study parameter inference in an homogenous population, then we repeat the analysis for heterogenous populations. We simulated 100 datasets for the homogeneous population study and 20 datasets for the heterogeneous population study, using known parameters, and fitted the model to each dataset. The dataset on which a model is fitted is referred to as the training set; we then validate performance on the other (independent) datasets. Since the time of death is stochastic, the OS curves are subject to noise, resulting in variation in the estimated parameters (and optimal solutions) across the datasets.

For homogenous populations the model predictions are in good agreement with the data Figure 11. The MLEs successfully recover the parameters *µ*_1,1_ and *µ*_2,1_ (Figure 11 **C, D**); however the dynamic parameters *d* and *g* have higher variation across data sets, Figure 11 **E, F**. The ratio of the standard deviation of MLEs to the true values are: 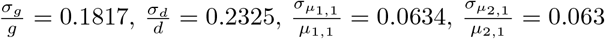. In fact the MLEs of *g, d* were highly correlated, Figure 11 **H**, suggesting a strong (linear) dependence between *g, d* (correlation coefficient of 0.9945).

**Figure 11.**
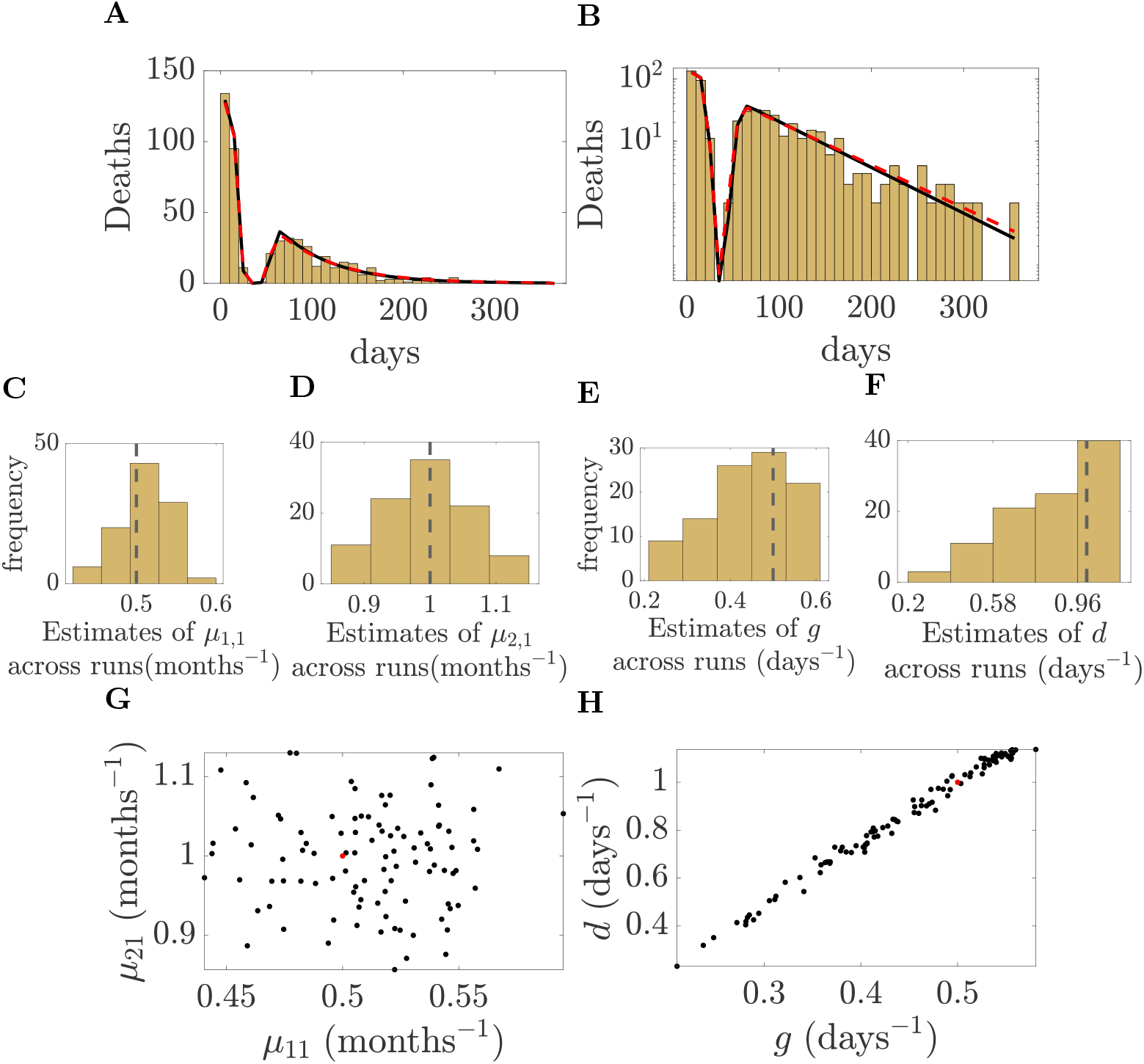
Parameter estimation from the survival curve of a homogeneous population. **A–B**: Linear and log scales of 10-day binned survival data for a single dataset (500 patients under 20 day MTD treatment). OS curve with true model parameters shown (red dashed) and fitted model (black). **C–F**: Histograms of maximum likelihood estimates for *µ*_1,1_, *µ*_2,1_, *g*, and *d* for 100 (independent) simulated datasets; dashed line shows the true values. **G/H** Scatter plots of maximum likelihood estimates for the 100 datasets for **G** mortality rates *µ*_11_, *µ*_21_, and **H** tumour growth parameters *d, g*. Red dot shows true values. Parameters are MLE and performed separately for each of 100 simulated homogeneous population datasets of 500 patients each. True parameter values: *µ*_1,1_ = 1*/*2 months^−1^, *µ*_2,1_ = 1 month^−1^, *g* = 0.5, days^−1^, *d* = 1, days^−1^. The initial tumour size is *N*_0_ = 10^6^ cells and the carrying capacity is *K* = 1.297 × 10^9^.

From the estimated parameters we can compute the optimal treatment duration for each dataset. The period of optimal drug administration varied, mean 38.8 days, standard deviation 11 days, which compares to 31.6 days for the optimal duration with the true parameter values, Figure 12-**A**. This variation was a consequence of the large variation in the MLEs for *g, d*, with *g* − *d* having the highest correlation with MTD duration, Supplement J. In fact there was a strong trend between the optimal MTD duration with the estimated *g* − *d*, Figure 12-**B**. The optimal treatments based on model parameters estimated from OS curves improved survival relative to the original 20-day MTD therapy, Figure 12C. This is evident from the OS curves where the number of relapsing patients is drastically reduced Figure 12**C**. Despite the variation in the MTD duration, the OS curves are very similar, and give a performance close to that of the optimal treatment duration based on true parameters Figure 12 **D**. Since the populations are homogeneous, the optimal solution estimated on the training data had statistically identical performance to those estimated on independent (validation) datasets, Figure 12 **C**. The longer MTD duration of the optimal solutions compared to 20 day MTD resulted in higher mortality during treatment through TRM, but the substantial gains in survival post-treatment (reduced relapse frequency) gave a higher survival probability, Figure 12**E-G**. Thus, estimated optimal solutions substantially improve overall survival on homogenous populations.

**Figure 12.**
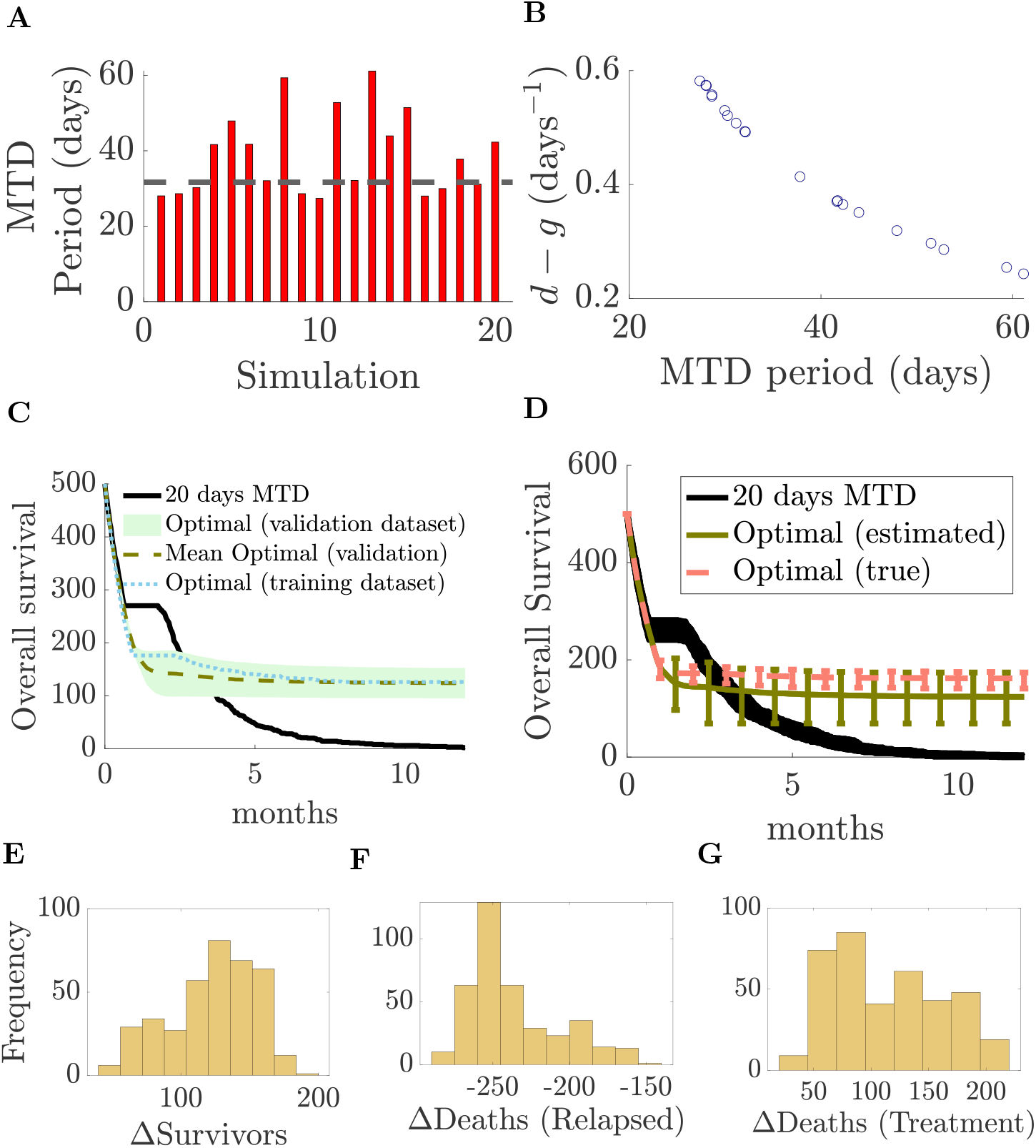
Performance of estimated optimal solutions on homogeneous populations. **A.** Bar plots of the optimal drug administration period for 20 simulated datasets. The black horizontal line indicates the optimum based on true parameter values. **B**. Scatter plot of the estimated tumour net decay rate (*d* − *g*) versus the optimal MTD administration period using estimated parameters for 20 datasets. **C**. Overall survival curves for a single dataset of 500 patients comparing the original 20-day MTD treatment (black, the fitting data), the optimal treatment for parameters estimated on that (training) dataset (dotted), and survival curves for optimal treatment for parameters from the 19 independent (validation) datasets. The green shaded region represents the min-max envelope across the 19 validation datasets, and the green dashed line shows the mean. **D**. Overall survival curves across all 20 datasets of 500 patients comparing the original 20-day MTD treatment (black), validation curves under the optimal treatment using parameters estimated from the other 19 independent datasets (green: min-max envelope with mean curve), and the optimal treatment based on true parameter values (pink: min-max envelope with mean curve). **E–F**. Histograms of differences (optimal minus 20-day MTD) for **E** total survivors, **F** post-treatment deaths (relapse mortality), and **G** deaths during treatment in the validation datasets (20 estimated optimal therapies evaluated on 19 independent datasets). Datasets as Figure 11.

To examine performance under population heterogeneity we generated simulated datasets (500 patients, 20-day MTD) with normally distributed model parameters, mean 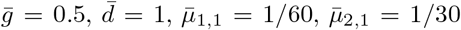, and a coefficient of variation of 10%-25%. We repeated the analysis above. For heterogeneous populations with a coefficient of variation CV=10%, there was an excellent reproduction of the training set survival curve, Figure 13**A-B**, and the fitted parameters were in good agreement with the data, the MLEs lying within the distribution of patient parameters Figure 13**C-F**. Histograms of the estimated parameter values across 20 independent datasets are shown in Figure 14**A-D**, with excellent recovery of the true mortality rates. There is a bias in MLEs for *g, d*, both *g, d* being overestimated (Figure 14**A**,**B**), whilst a strong linear relationship between the MLEs of *g, d* was apparent with slope of 1 (Figure 14**F**). Thus, the estimated tumour decay rate (*d* − *g*) was approximately constant across datasets and had little variation between simulated datasets, Figure 14**E**. Of the twenty simulated datasets, all but one dataset had an estimated *µ*_1,1_ and *µ*_2,1_ that lay within the 68% level set of the source Gaussian (Fig. 14**G**), whereas most inferred *g* and *d* values fell outside the 95% region (Fig. 14**F**). Bias in MLEs is not unusual for finite sample sizes.

**Figure 13.**
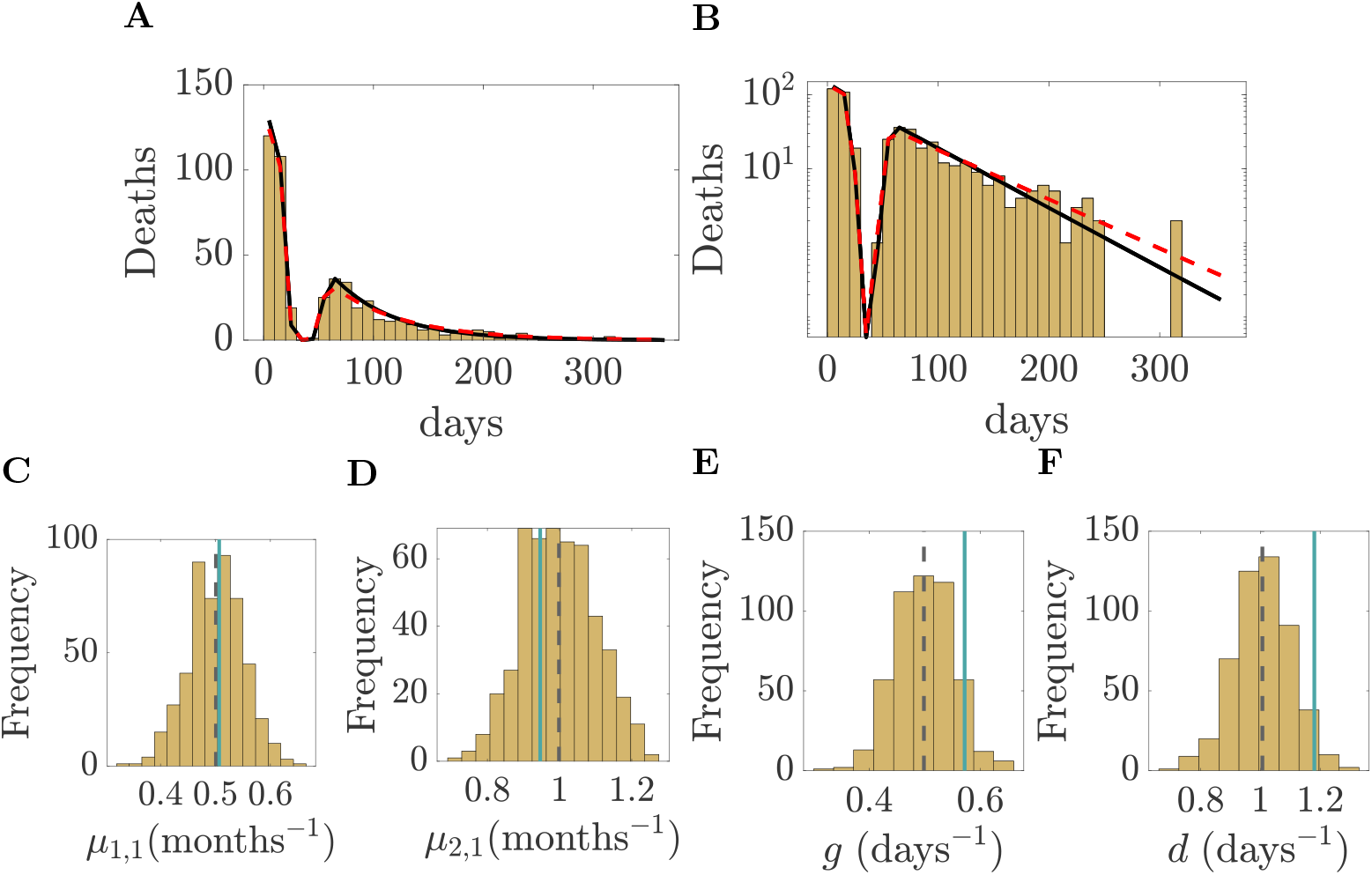
Parameter estimation from survival curves of a heterogeneous population with coefficient of variation CV= 10%. **A–B**: Linear and log scales of OS curves for a single dataset (500 patients, 20-day MTD, 10-day bins). OS curve from a model with true mean parameters shown (red dashed) and the fitted model (black). **C–F**: Histograms of patient parameters for dataset in **A**,**B**; black dashed lines are means of source (Gaussian) distribution, and green lines are the maximum likelihood estimates for *µ*_1,1_, *µ*_2,1_, *g*, and *d*. 500 patients are simulated with parameters drawn from a Gaussian distribution, means 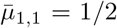 months^−1^, 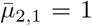 months^−1^, 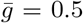 days^−1^, 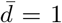 days^−1^, and a coefficient of variation 10%. The initial tumour size is *N*_0_ = 10^6^ cells and the carrying capacity is *K* = 1.297 × 10^9^.

**Figure 14.**
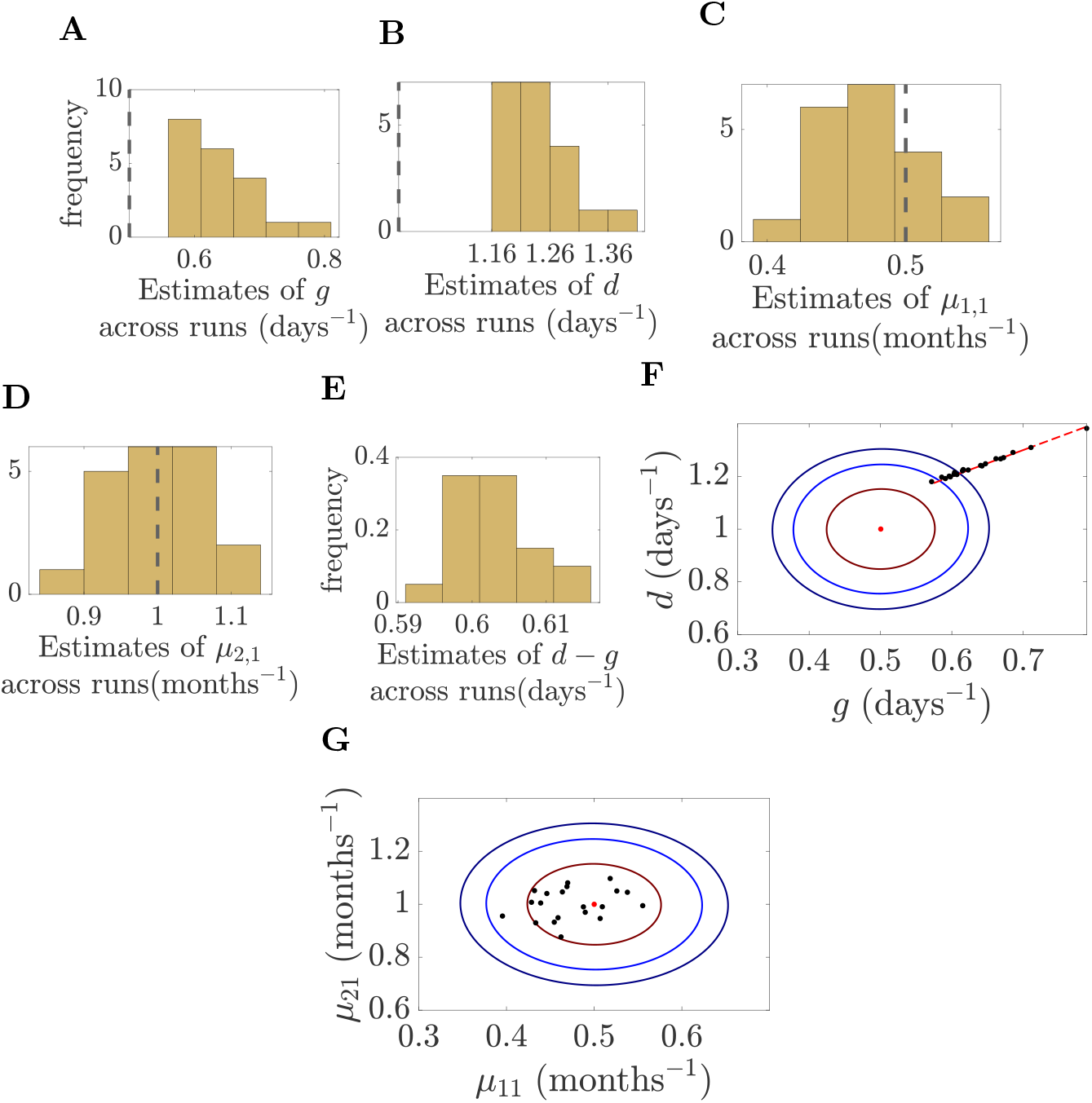
Parameter MLEs for heterogenous populations with coefficient of variation 10%. **A-E.** Maximum likelihood estimate histograms for *µ*_1,1_, *µ*_2,1_, *g, d* and *d* − *g* on 20 datasets; dashed lines indicate the source (Gaussian) distribution means (the true value in **E** is *d* − *g* = 0.5 day^−1^ is omitted due to scale). **F/G**. Scatter plots of maximum likelihood estimates for **F** *µ*_1,1_, *µ*_2,1_ and **G** *g, d*. Red dots indicate the source distribution population means; dark red, light blue and dark blue contours show the 68%, 95%, and 99% level sets of source distribution. In **G**, the red line (parallel to *d* = *g*) highlights that *d g* remains nearly constant across MLEs. Parameters are estimated on simulated survival curves under a 20 day MTD treatment regimen on 500 patients with model parameters drawn from Gaussian distributions with means 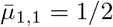 months^−1^, 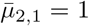 months^−1^, 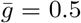 days^−1^, 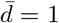 days^−1^, and coefficient of variation 10%. 20 independent datasets are simulated. The initial tumour size is *N*_0_ = 10^6^ cells and the carrying capacity is *K* = 1.297 × 10^9^.

Despite variability in the estimates of *g* and *d*, Figure 14**F** the optimal drug duration for each of the datasets (based on the parameter MLEs) was very consistent across the 20 independent datasets, with mean and standard deviation of 26.5 days, 0.2 days respectively, of continuous MTD (Figure 15A), a shorter MTD duration than the optimum 31.6 days MTD for the mean parameters of the source population variation Gaussian. This consistency is in contrast to the high variability observed in homogeneous populations, Figure 12**A**. The MTD duration has a strong anti-correlation with *d* − *g* (Figure 15B), (*r* = −0.82, Supplement J), and shows moderate correlation with the mortality rates *µ*_1,1_ and *µ*_2,1_ (|correlation| *>* 0.39, Supplement J) in contrast to the homogeneous case where correlations were small, Supplement J. However, the MLE of *d* − *g* shows little variability (Figure 14**F**), and both mortality rates are well estimated, lying within the 68% level set of the source Gaussian (Figure 14**G**). This lack variation explains the consistent optimal solution durations across datasets; the correlation with *g* + *d* is weak (|correlation| *<* 0.39, Supplement J), which helps explain the consistency of the optimal MTD.

**Figure 15.**
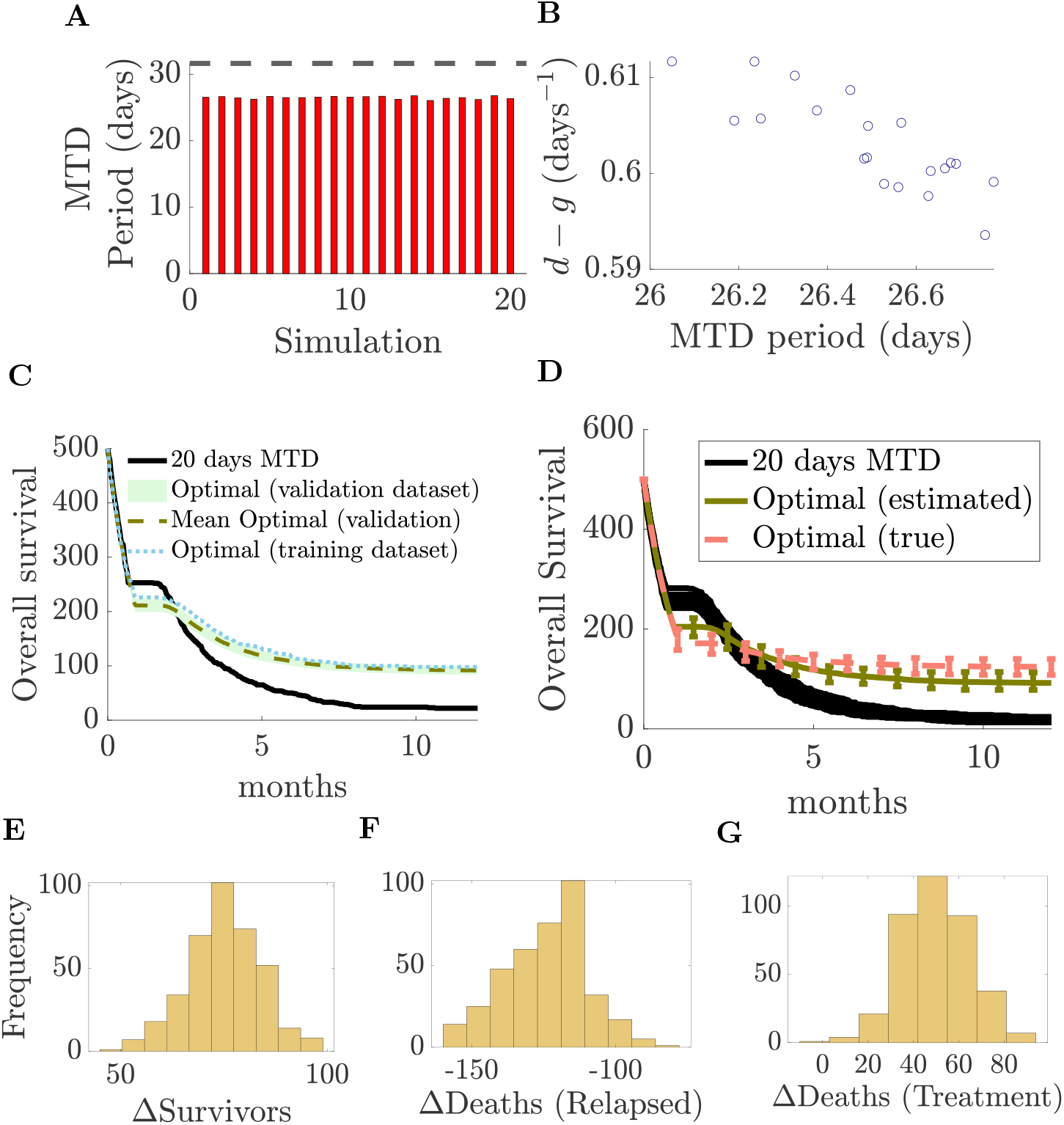
Performance of optimal therapies based on estimated parameters for heterogenous populations (CV 10%). **A.** Optimal drug administration period based on parameters inferred from each simulated dataset (20 simulations, 20-day MTD). The horizontal line indicates the optimum using the source Gaussian mean parameters. **B**. Scatter plot of the estimated tumour net decay rate (*d* − *g*) versus the optimal MTD administration period using estimated parameters for 20 datasets. **C**. Overall survival curves for a single dataset of 500 patients comparing the original 20-day MTD treatment (black, the fitting data), the optimal treatment for parameters estimated on that (training) dataset (cyan dotted), and survival curves for optimal treatment based on parameters estimated from the 19 independent (validation) datasets (green, min-max envelope with mean). **D**. Overall survival curves across all 20 datasets of 500 patients comparing the original 20-day MTD treatment (black), survival curves under the optimal treatment based on parameters estimated from the 19 validation datasets (green, min-max envelope with mean), and the optimal treatment based on mean parameter values of the source Gaussian (pink, min-max envelope with mean). **E–F**. Histograms of differences (optimal treatment minus 20-day MTD) for **E** total survivors, **F** post-treatment deaths (relapse mortality), and **G** deaths during treatment in the validation datasets (20 estimated optimal therapies evaluated on 19 validation datasets). Datasets simulated as Figure 14.

The OS curves under the optimal treatment estimated on the training dataset demonstrates substantial improvement; for the dataset of Figure 15 **C** 126 survived compared to 2 under 20 days MTD. This improvement is in fact retained under the optimal therapy based on the parameters estimated from (independent) validation datasets, the training set and validation set survival curves being very similar, Figure 15 **C**. Across the 20 datasets, the OS curves consistently demonstrate substantial improvements over the 20-day MTD, Figure 15 **C**, with similar performance to the optimal therapy based on the mean parameters of the population variation source Gaussian. The increased OS observed for the estimated optimal solutions is again driven by a reduced relapse rate, despite a higher TRM (Fig. 15 **E-G**).

When population heterogeneity increased to CV = 25% (Figures 16–18), similar trends were observed to 10% heterogeneity. Model predictions remained in good agreement with the data (Fig. 16), with MLEs lying within the population variation and with excellent reproduction of the OS curve. All estimates of *µ*_1,1_ and *µ*_2,1_ lay within the 68% confidence region, whereas all estimates of *g* and *d* fell outside it (Figure 17). MLEs of *µ*_1,1_.*µ*_2,1_ lay close to the true values, whilst *g, d* were overestimated, Fig. 18. As previously, the tumour decay rate under MTD (*d* − *g*) remained approximately constant across simulations (Fig. 18 B).

**Figure 16.**
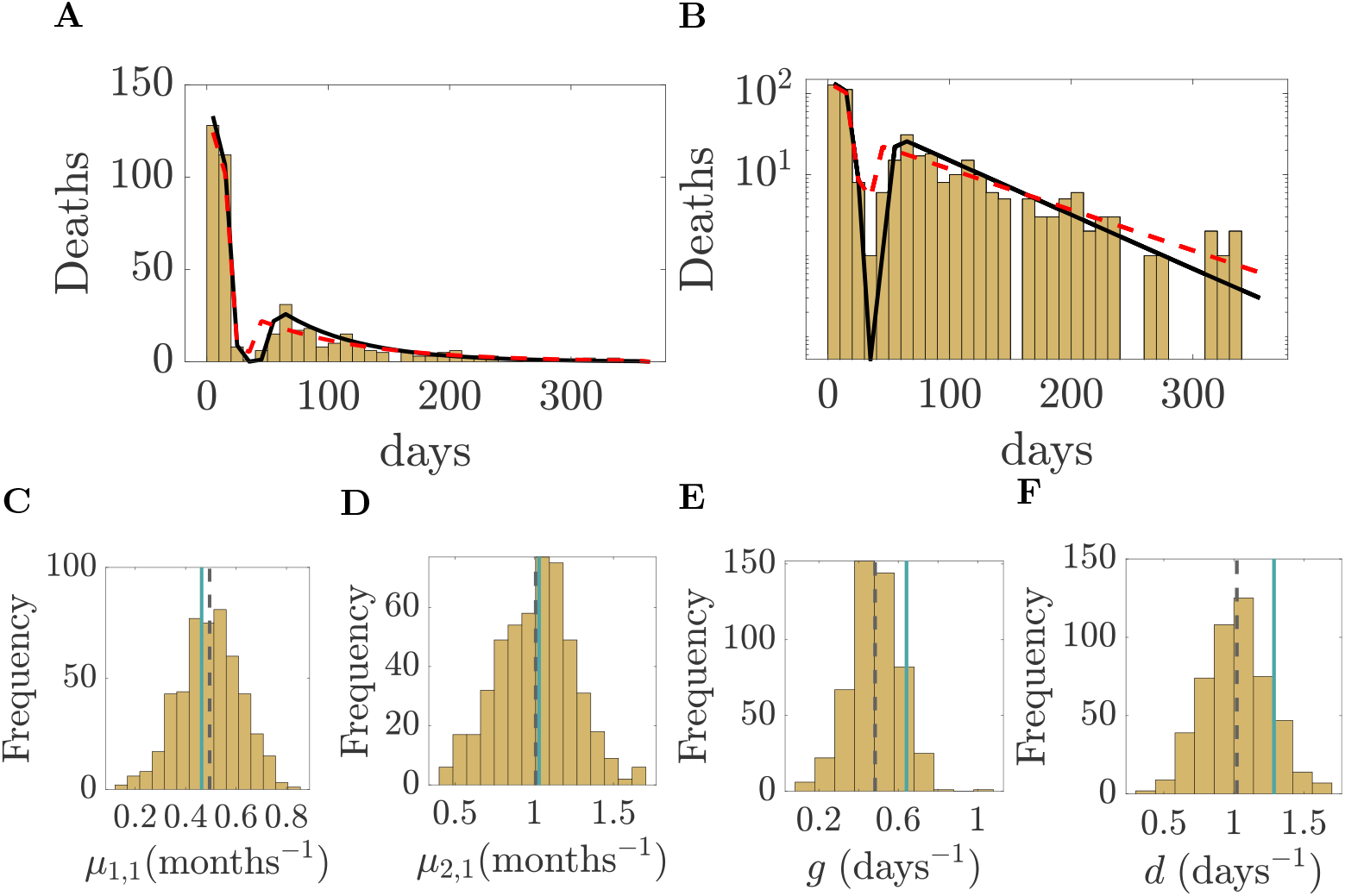
Parameter estimation from survival curves of a heterogeneous population with coefficient of variation CV= 25%. **A–B**: Linear and log scales of OS curves for a single dataset (500 patients, 20-day MTD, 10-day bins). OS curve from a model with true mean parameters shown (red dashed) and the fitted model (black). **C–F**: Histograms of patient parameters for dataset in **A**,**B**; black dashed lines are means of source (Gaussian) distribution, and green lines are the maximum likelihood estimates for *µ*_1,1_, *µ*_2,1_, *g*, and *d*. 500 patients are simulated with parameters drawn from a Gaussian distribution, means 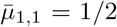 months^−1^, 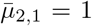 months^−1^, 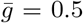 days^−1^, 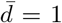 days^−1^, and a coefficient of variation 25%. The initial tumour size is *N*_0_ = 10^6^ cells and the carrying capacity is *K* = 1.297 × 10^9^.

**Figure 17.**
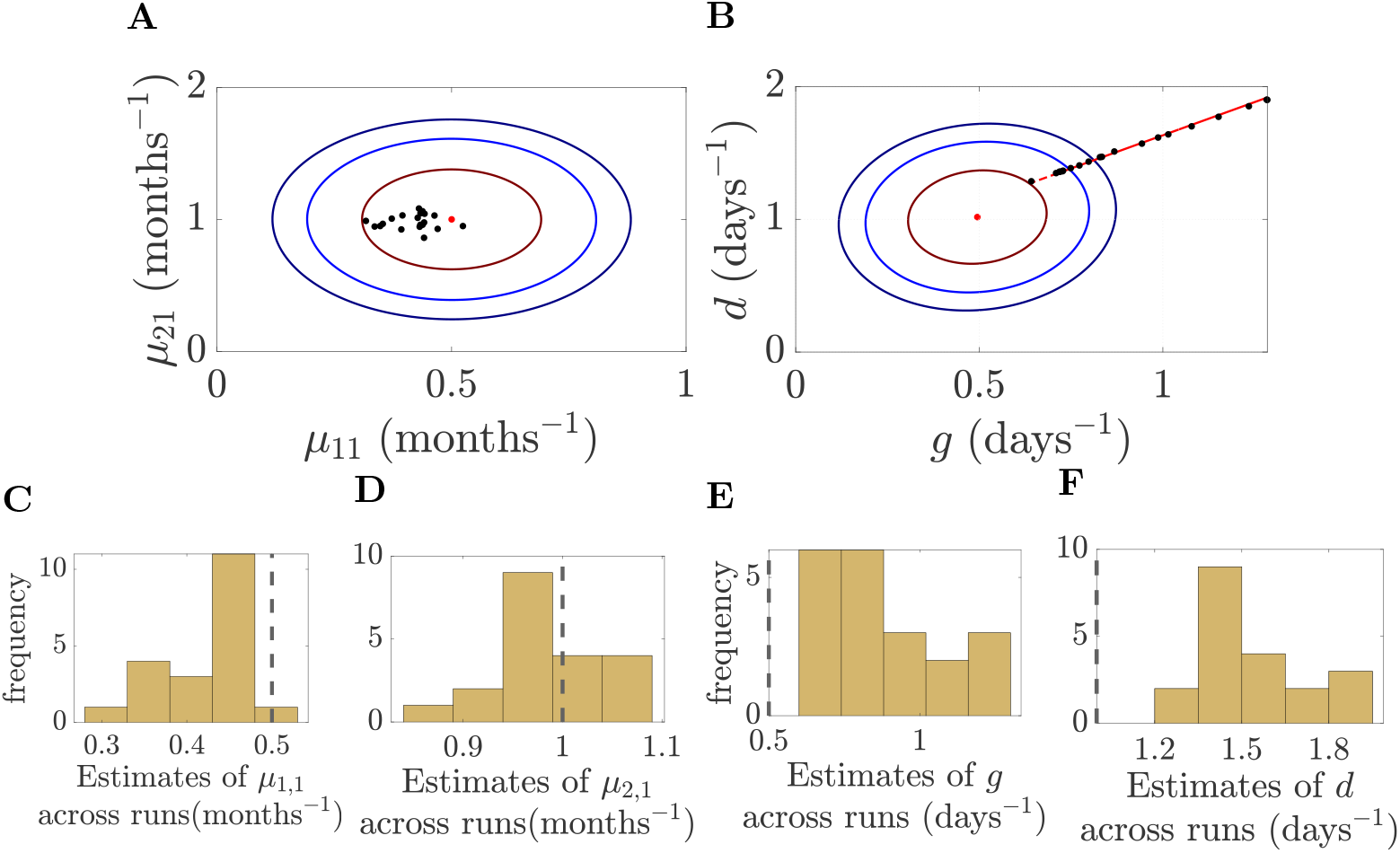
Inferred values of **A** *µ*_1,1_, *µ*_2,1_ and **B** *g, d* across all survival curves (20 simulations; parameters normally distributed with means 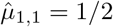 months^−1^, 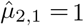 months^−1^, 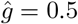 days^−1^, 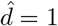 days^−1^, and coefficient of variation 25%). The initial tumour size is *N*_0_ = 10^6^ cells and the carrying capacity is *K* = 1.297 × 10^9^. Red dots indicate the true population means; red, light blue and dark blue contours show the 68%, 95%, and 99% confidence regions. In **B**, the red line (parallel to *d* = *g*) highlights that *d* − *g* remains nearly constant across estimations. **C**–**F**: Distributions of inferred *µ*_1,1_, *µ*_2,1_, *g*, and *d*; dashed lines indicate the true means.

**Figure 18.**
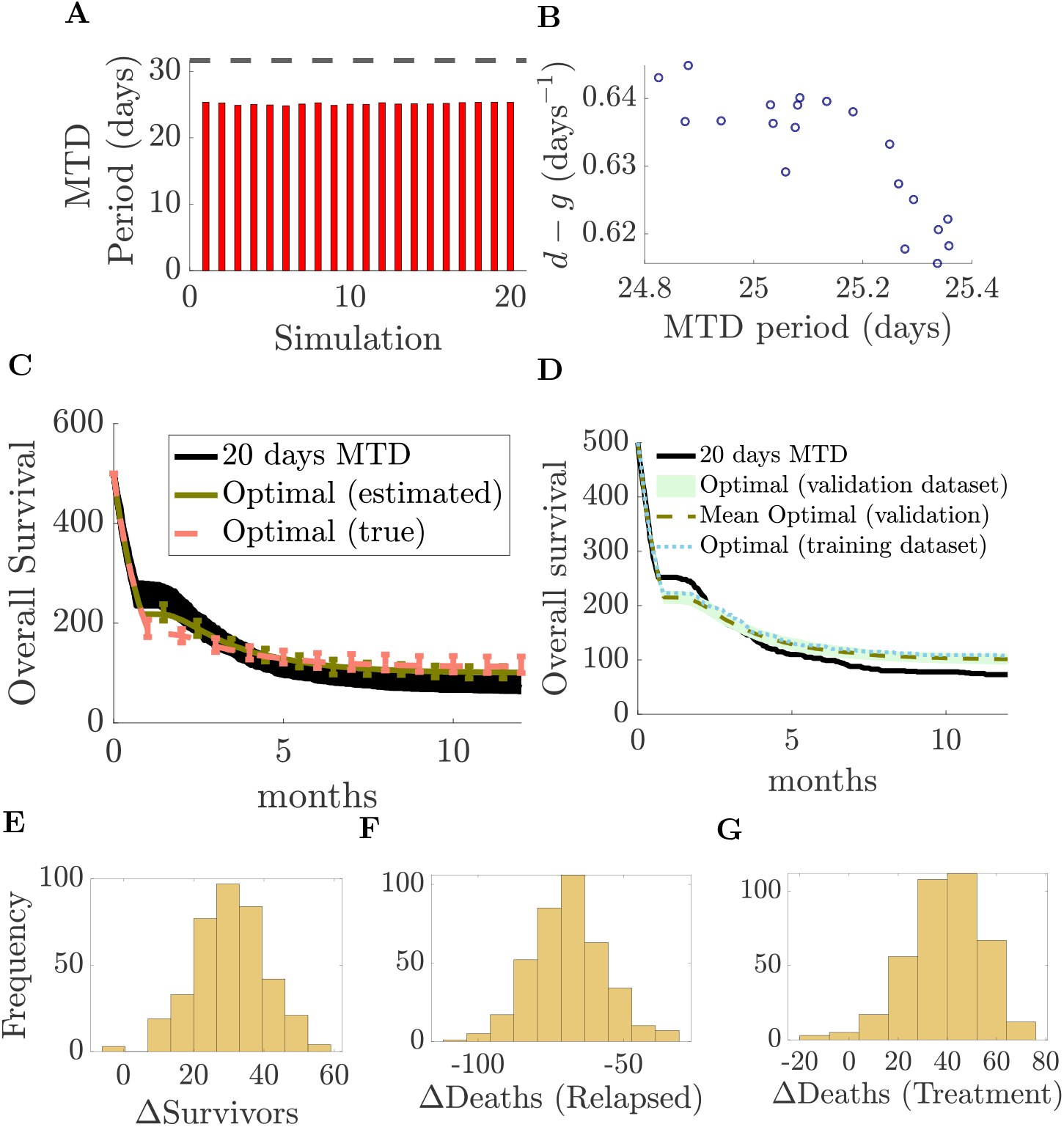
A Optimal drug administration period based on parameters inferred from each simulated dataset (20 simulations; parameters normally distributed with means 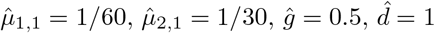, and coefficient of variation 25%). The initial tumour size is *N*_0_ = 10^6^ cells and the carrying capacity is *K* = 1.297 × 10^9^. The horizontal line indicates the optimum using the population mean as effective parameter values. B Tumour decay rate under MTD (*d ™ g*) from inferred *g* and *d* for each simulation; the horizontal line shows the average *d ™ g* across the population. C Comparison of optimal treatments with 20-day MTD in terms of overall survival. Pink lines show survival curves from 100 simulations of 500 patients under an optimal solution in A (using the estimated parameters of one simulation), with the green line denoting the mean survival. The red line shows the mean survival under the optimal treatment using the population mean parameters from A. D–F Histograms of differences in number of survivors, relapse mortality, and treatment mortality between the optimal treatment (estimated parameters) and 20-day MTD.

Estimated optimal solutions were consistent across datasets with around 25 days of continuous MTD (Fig. 18 A). The improvements of the estimated optimal solutions over 20-day MTD were less substantial at 25% heterogeneity, Fig. 18 **C-D**, but similar to the optimal therapy based on mean population parameters. Population heterogeneity in fact improves the overall survival of our base-line 20-day MTD, so possible gains are reduced. This highlights that uniform treatment strategies are less effective under high population heterogeneity, emphasisng the need for population stratification and personalised therapy. As before improvements in OS were again due to lower relapse rates Fig. 18 **E-G**.

## 7 Conclusions

Here we developed CLP, the expected patient lifetime, as a tractable objective for cancer therapy optimisation within a Poisson process framework, allowing incorporation of key clinical events that impact patient lifetime such as patient death and tumour elimination. The harmful and beneficial impacts of treatment are quantified through the change of these event rates, and the net benefit of therapy is then naturally determined by the combined effect on the expected lifetime. By evaluating event probabilities the CLP classifies post-treatment patient outcome and provides associated outcome probabilities. Key factors that influence patient survival (such as tumour size, tumour eradication time, and patient age) are incorporated directly into the objective functional. This framework offers a novel perspective on the formulation of clinically relevant OCT problems that have a direct link to clinical endpoints and treatment performance criteria (such as overall survival (OS) curves) rather than relying on qualitative and subjective measures of drug toxicity or tumour reduction. Importantly, it provides insights into the development of OCT objective functions that can in principle be calibrated from real-world, patient-specific data.

A major challenge in cancer therapy optimisation is quantifying anti-tumour efficacy for incorporation into an objective for accurate treatment comparisons. Common metrics [1, 20, 31, 44], such as tumour size at the end of treatment and average tumour size throughout the treatment period, fail to capture long-term/post-treatment effects - in deterministic models, tumours will regrow after therapy (a 100% relapse rate) and failing to allow for this can lead to medically ill advised optimal solutions such as delayed treatment [42, 43, 45]. The CLP, and our earlier LEP framework [69], address this by incorporating the probability of tumour elimination (PTE) into the objective function. The PTE captures the likelihood of successful tumour clearance, accounting for stochastic die-out events that can occur even after the end of therapy. The CLP naturally allows other stochastic events to be included as Poisson processes, although dependence of the event rate on state variables is needed. We constructed a CLP with patient mortality events from 3 causes: treatment-related mortality (TRM), mortality due to tumour progression and age-related mortality. Consequently, the probability of patient survival depends on tumour progression, drug scheduling and patient age. The CLP formulation is flexible, not being restricted to any specific functional forms for drug or tumour-related mortality rates, and other events such as metastasis and mutation can be included.

The CLP has a number of fundamental properties. Firstly, there is no subjective weighting of competing objectives as is typical of OCT applications to cancer therapy. The competing objectives of anti-tumour efficacy and chemotherapy toxicity are implicitly weighted by their impact on patient expected lifetime. Secondly, provided drug application has ended by the control horizon time *T*_0_, the payoff and the optimal solution are independent of *T*_0_. This follows since the CLP only depends on *T*_0_ through the constraint on *u*; thus *T*_0_ can be decreased without changing the CLP and the optimal solution if the optimal solution has *u* = 0 prior to *T*_0_. Thus, provided the horizon is sufficiently long, there is a single optimal solution for a given initial tumour size independent of *T*_0_. Thirdly, the CLP naturally categorises outcomes with associated outcome probabilities. It shares all these properties with the LEP [69].

To illustrate CLP optimisation, we used a single-compartment logistic tumour growth model with a tumour-dependent mortality rate that was linear in tumour size, and a TRM rate linear in the drug dose. The optimal control problem was solved using a classical direct numerical method that discretizes the control as a piecewise constant function, optimizing both drug concentration and the discretization mesh, Supplement G. We only observed bang-bang optimal therapies, in fact we had 3 types of solution: MTD for the duration, no treatment and treat-and-stop solutions with drug administered continuously at MTD from *t* = 0 up to a specific (optimal) time point *x*. MTD for the duration was a result of an insufficiently long control horizon *T*_0_, thus there are only 2 optimal strategies for sufficiently long control horizons *T*_0_. This divides patients into treatable and untreatable groups. Administering MTD for less than the optimal duration substantially reduced the PTE (Figure 3 **C**), while extending treatment beyond the optimal period increases TRM with little additional PTE gains. For untreatable patients, the risk of TRM outweighs any potential gain in PTE, rendering treatment ineffective. Treatability depends on patient-specific characteristics, in particular a critical drug tolerability threshold separates treatable from untreatable patients (Figure 8), increasing strictly with tumour size; larger tumours require more drug to achieve comparable tumour reduction and a similar PTE (Figure 7). Maximizing life expectancy for patients near this boundary may involve high TRM risk; our simulations show that even patients with high drug sensitivity (probability of drug-related death ¿0.9) can gain a net average survival benefit. Optimal solutions depend little on age (Figures 6 and 4), with 20- and 90-year-olds (with identical drug tolerability) sharing nearly identical drug administration windows for the same tumour size. However drug tolerability likely decreases with age, which would then differentiate treatment and outcomes by age since drug administration periods change with drug tolerability Figure 7**A**. Overall, the optimal treatment duration is primarily driven by tumour size whilst PTE is set by patient drug tolerance: the greater the tolerance to drug toxicity, the more extended the optimal therapy which increases PTE.

The CLP, being based on clinical events and biological processes, is in principle parametrisable from data. For instance age-related mortality rates can be inferred from life-tables, Figure 1. The continuous stochastic process modelling framework of the CLP naturally generates survival curves (Figure 4), thus providing direct comparison to, and enabling model parametrisation from clinical data. Thus, clinically relevant endpoints such as the proportion of patients who die from treatment, relapse, or are cured can be used to constrain the parameters. We illustrated parameter estimation from overall survival curves using simulated data, simulating a dataset of 500 patients. We estimated 4 parameters from OS curves using maximum likelihood estimation which enabled optimal drug administration duration to be estimated. Inference of the mortality rates *µ*_1,1_, *µ*_2,1_ was excellent and robust against patient heterogeneity. Tumour growth rate estimates showed greater variability although the net growth rate *d* − *g* under MTD was highly constrained, i.e. 3 parameters could be estimated from OS curves (the net growth rate *d* − *g* was biased). Cancer survival curves are in fact well fitted by the 2 parameter Weibull distribution, and 3 parameters when there is a cure fraction, [56]. The estimated optimal solutions improved survival and were robust against patient heterogeneity, although the optimal treatment gains deteriorated as patient heterogeneity increased. Optimal solutions had higher TRM, but that is compensated for by lower relapse rates. Our analysis of model inference from OS data emphasised the need for tackling patient heterogeneity, since optimal solutions are optimal for a defined set of parameters and thus performance deteriorates on under population variation. Structuring by tumour type and patient drug tolerance (possibly related to age and health) is thus needed, or personalised treatments determined, for instance under adaptive therapy with updating of parameter estimates on subsequent measurements. In practice survival data is typically censored, which can be incorporated into the inference but will reduce the information in the dataset.

Inference of model parameters and model identifiability depends on data quality and whether the data is informative on the parameters. Overall survival curves contain crucial information, but likely these need to be supplemented with additional data to infer all the model parameters, particularly as model complexity increases. There are a number of possibilities. More information on patient progress would be valuable (individual participant data), for instance cause of death data could distinguish TRM, tumour-induced death and death from unrelated causes, whilst the time when the disease is undetectable (below detection threshold) may be informative on tumour dynamics. The size of solid tumours can be monitored by ultrasound, MRI or CT scans, whilst in some cancers, tumour progression can be monitored longitudinally from blood samples, for instance in leukemias (liquid cancers) the leukemic cell count can be measured [70], whilst in prostate cancer and ovarian cancer the prostate specific antigen PSA [41, 78] and serum cancer antigen-125 (CA-125) [4,11] respectively can be monitored. Circulating tumour cells (CTC) [33, 66] is an emerging method but resolution is low. All of these methods have interpretation issues, [60] and likely need validated dynamic models to enable their use in quantification analyses, but in principle such data could be used to infer tumour dynamic parameters within specific cancer type models, [39].

The CLP formulation provides a general framework for constructing objectives for evaluating clinical therapy. Key ingredients are a tumour growth dynamics model, models for drug action/efficiency on the tumour and impact on the patient, and key event rates and their dependencies, such as mortality. A major cause of death associated with several cytotoxic drugs is chemotherapy-induced myelosuppression [13], a mechanism that could be incorporated using haematopoietic models [12, 21]. Other common causes of TRM include organ damage [80] as well as neurological [72] and cardiovascular disorders [52]. Models of these processes could be included, eg cardiovascular toxicity [62], replacing the mortality rate *µ*_2_(*N*) with a mechanistic model. Generally, the CLP can be extended to multi-compartment models that capture tumour heterogeneity, for instance tumours with (not exhaustive) chemo-sensitive and chemo-resistant cell subpopulations [34, 54] or continuous drug sensitivity variation [57], multiple genetic subclones within a tumour [65], cell cycle states [42, 61], immune-tumour models [47] and models that include normal tissue compartments [14, 26, 30, 63, 82], as suggested above. There are also a number of mathematical models for specific cancers that accurately reproduce tumour growth under treatment [16, 23, 28, 32]. Additional complexities include drug pharmacokinetics/dynamics, a dependence on patient health including performance status [75], whilst patient mortality can arise from multiple correlated causes, [6] and treatment can lead to secondary AML [25]. Quality of life may also be an important consideration. A range of (validated) predictive models are emerging that could be incorporated within a CLP framework, both from analysis of clinical trial data [17, 68, 77] and national databases such as CPRD, including for instance the impact of comorbidity history on mortality, [55] which naturally leads to personalised therapy. Psychological factors [49] and patient discomfort [40], which may significantly influence clinical decision making, remain difficult to quantify, although likely of more importance for patients near the untreatable boundary. Key challenges in such extensions is firstly model parametrisation, and secondly computing the tumour elimination probabilities to reflect the joint eradication of all cancer cell subtypes. These will be the subject of future work.

## Data Availability

All data produced in the present work are contained in the manuscript

## Acknowledgment

BDET was supported by the EPSRC Postdoctoral Pathway Fellowship scheme (University of Warwick, grant number EP/Z535011/1) and was affiliated with the Mathematics of Systems (MathSys) CDT for part of this work.

## Supplements

### A Asymptotic behaviour of the TCP

The TCP is the probability that the tumour gets eliminated by time *t*; thus taking the limit of the TCP to infinity gives the probability that the tumour eventually gets eliminated by some point during or after the treatment. Naturally, the TCP is an increasing function with time as stochastic elimination event that can occur during or even after the treatment increase the likelihood of tumour elimination; this can be also verified by differentiationg Equation **??**. In this Section we show that when the drug free mortality rate 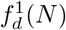 is zero, the TCP is strictly increasing and eventually asymptotes to 1 regardless of the treatment. In the case where 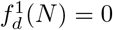

#### Theorem A.1

*In the case where f*_*d*_(*N*, 0) = 0 *the* TCP *remains constant in the absence of drug, whereas in the case where f*_*d*_(*N*, 0) *>* 0 *the* TCP *increases and asymptotes to one*.

If *f*_*d*_(*N*, 0) = 0 and there is no drug administered in some interval *I* then

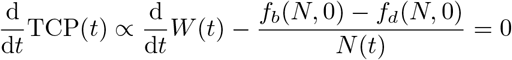

for all *t* ∈ *I*. In the case where *f*_*d*_(*N*, 0) *>* 0 the TCP strictly increases in *I* as 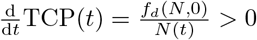.

For the asymptotic behaviour of TCP in the general case we have

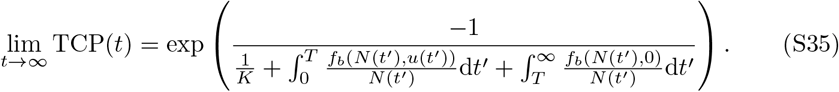

In the case where *f*_*d*_(*N*, 0) = 0, then 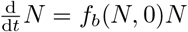 in the absence of drug. Thus

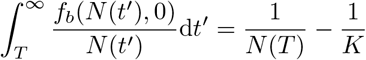

and so

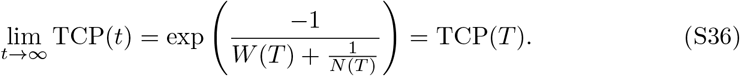

If *f*_*d*_(*N*, 0) *>* 0 then the limit (Equation S35) is one. Indeed, in that case we have

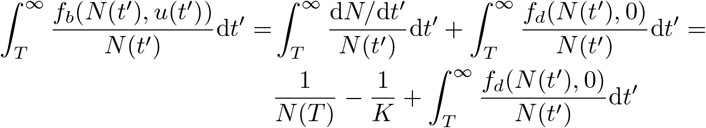

which gives:

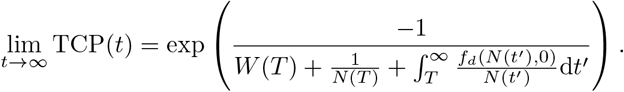

The death rate increases with *N* and approaches the birth rate once the tumour is in a neighbourhood of the carrying capacity. Consequently for some *t*_1_ *> T* with the property *N*(*t*) *> K* − *ε*, ∀ *t > t*_1_, *ε >* 0 and small:

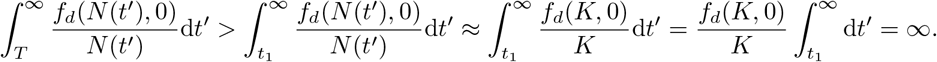

Hence, lim_*t*→∞_ TCP(*t*) = 1.

## B. Outcome of stochastic fluctuations when the average tumour size is above *ζK*

In this section we show that when the tumour size is close to the carrying capacity (i.e. *N*(*t*) ≥ *ζK* with *ζ* ≈ 1), the BP mortality does not influence the TCP as the latter is practically invariant. In general, the larger the average tumour size, the smaller the effect of stochastic cancer cell die out events. In the case where the tumour size is small stochastic fluctuations of the cancer cell population could effectively increase the likelihood of the tumour elimination.

Consider the interval *I* defined as *N*(*t*) ≥ *ζK*, ∀*t* ∈ *I*. Let *t*^*^ = min(*I*), with the property *N*(*t*^*^) = *ζK* and *t* an element of *I*. Then by definition:

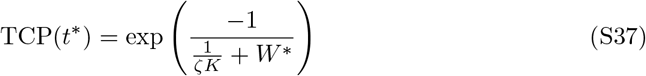

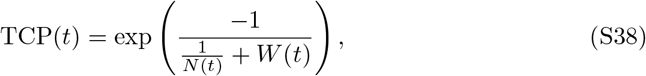

where *W* ^*^ = *W* (*t*^*^). Then

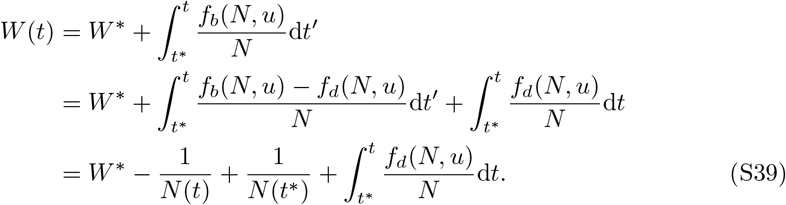

Equation S39 was derived by using the tumour dynamics

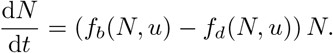

Since *f*_*d*_ is strictly increasing with *N* (Section 2.1), and *N*(*t*) ≥ *N*(*t*^*^) for all *t* ∈ *I*,

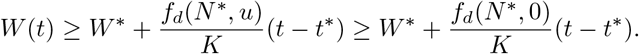

Then from Equation S38 we get

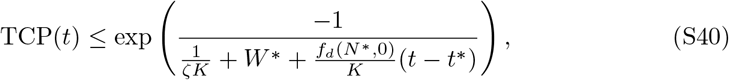

The carrying capacity is in the order of 10^9^ cells. Because 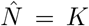 is a drug free equilibrium of the tumour size, *f*_*b*_(*ζK*, 0) = *f*_*d*_(*ζK*, 0) with the per-capita cancer cell growth rate *f*_*b*_(*N*, 0) in the order of 10^−1^ cells/day; thus the term 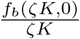 in Equation S40 is in the order of 10^−10^.

For example if (*t* − *t*^*^) = 100 years, then

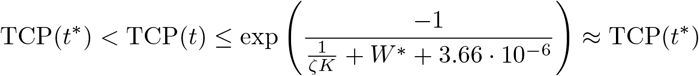

As a result, when *N ≥ ζK* over finite time scales, the BP mortality has little impact on the TCP, and drug-independent mortality does not meaningfully contribute to spontaneous tumour elimination. In contrast, drug-related mortality can reduce the average tumour size (*N*(*t*)) to levels where stochastic fluctuations significantly affect the elimination probability.

## C Derivation and Parametrisation of the Age related Survival Probability

Consider a random variable *A*, which expresses the age at which an individual dies. We estimate the probability that an individual of age *a* dies at age *a* + 1 :

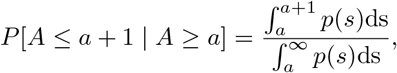

where *p*(*t*) is the PDF of *X*. Here, we have conditioned on the patient being alive at age *a* and we assume that mortality events follow a Gompertz Makeham distribution. The CDF is 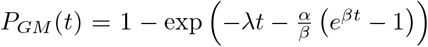 which gives the mortality probability *P*[*A* ≤ *a* + 1 | *A* ≥ *a*]:

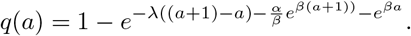

We calibrate the parameters *α, β* and *λ* of the age related mortality probability to national life tables from the [53]. The dataset provides historical mortality rates per 100,000 individuals, reported separately for women and men in the England, covering the period from 1981 to 2020. Each data point *q*_*a*_ represents the probability that an individual aged *a* in a given three-year period (e.g., 1981–1983, …, 2016–2018) dies before reaching age *a* + 1, Figure S19. The mortality distribution changes within each year; nevertheless to demonstrate the models behavior it is assumed that the age related mortality distribution is stationary and we assume that the death distribution is identical to that of the years 2016–2018. The data set that was used in this study contains the mortality rates of men in England that took place within the time period 2016-2018.

The parameters are estimated using a *χ*^2^ fit. In the dataset the rates *q*_*a*_ were given per *N* = 100000 people. We can thus assume that the parent population, at each bin is Poisson distributed with standard deviation 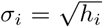, with *h*_*i*_ the number of people that died at age *a*_i_, i.e. *Nqa*. The error to the mean is then given by 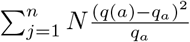 and we minimized:

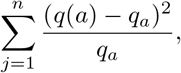

for *a* ∈ [20, 80]. The result of the fit is shown in Figure 1.

**Figure 19.**
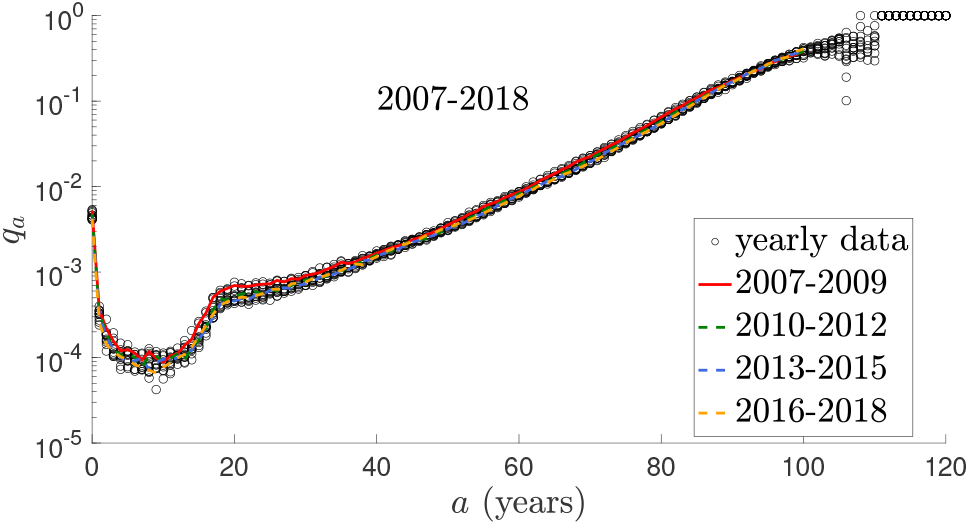
Historical mortality rates per 100,000 men in England within the years 2007 to 2018 as a function of the age. Black points represent mortality rates within each year. Red, green, blue and orange lines represent mortality rates within the time periods 2007-2009, 2010-2012, 2013-2015 and 2016-2018 respectively.

## D Details for the CEP derivation I

Using Equation 18 we get

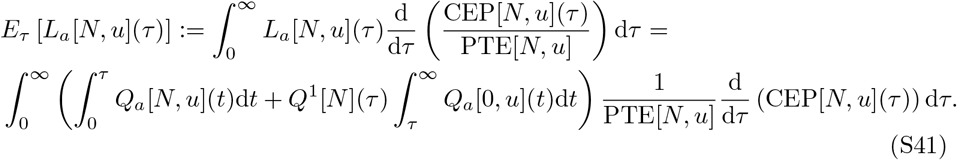

Notice that the integral 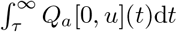 is finite, that lim_*τ*→∞_ *Q*^1^[*N*](*τ*) = 0 and that lim_*τ*→∞_ CEP[*N, u*](*τ*) = PLE[*N, u*]. Then applying integration by parts in Equation S41 yields

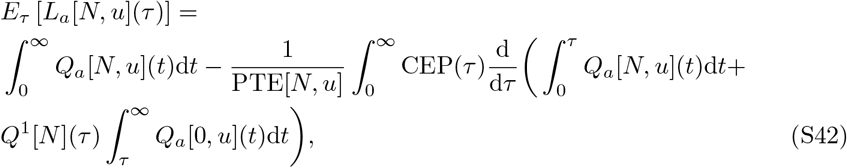

where for notational simplicity CEP[*N, u*](*τ*) is denoted as CEP(*τ*). Taking into account that 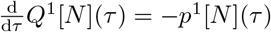 and that 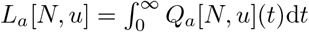 we get

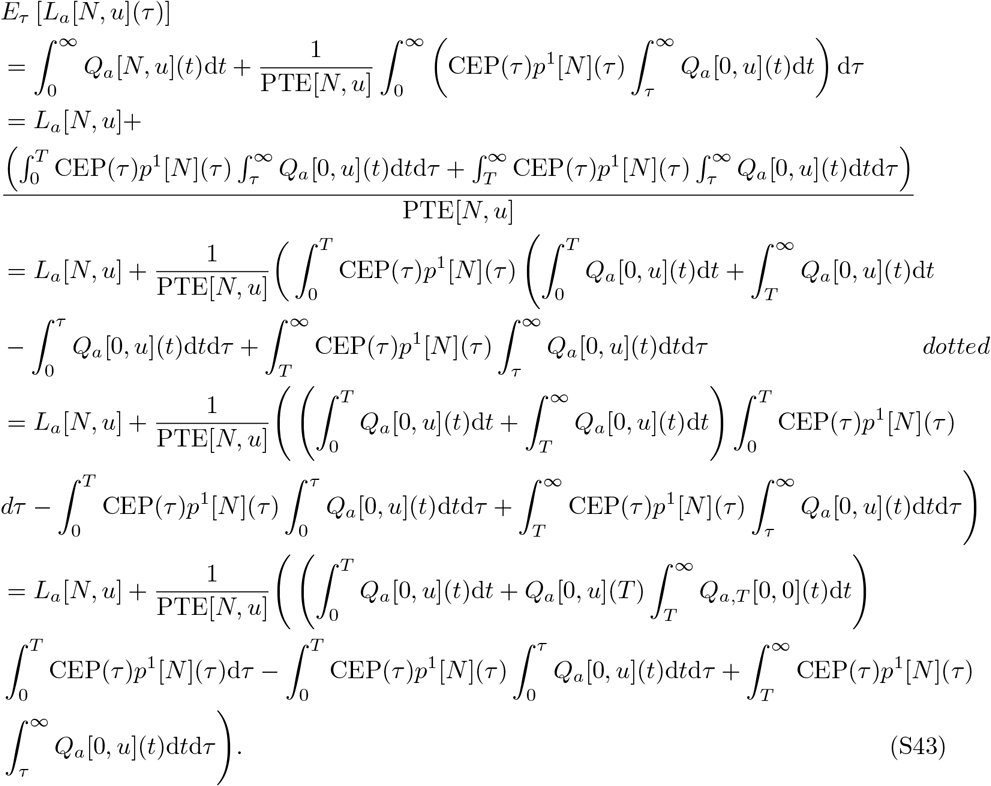

Combining Equations S43, 17 and 19 the CLP gets the form:

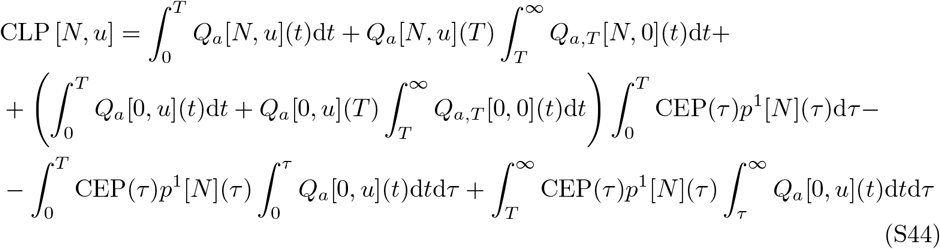

## E Analytical Derivation of *I*_0_ and *I*_1_ in closed form

In this Section we evaluate the integrals

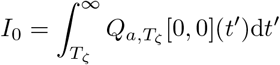

and

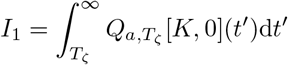

### Evaluating *I*_0_

By definition

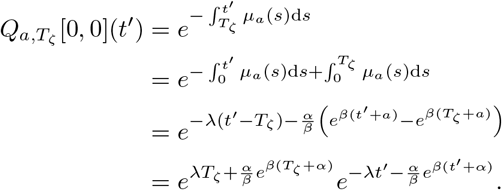

Then

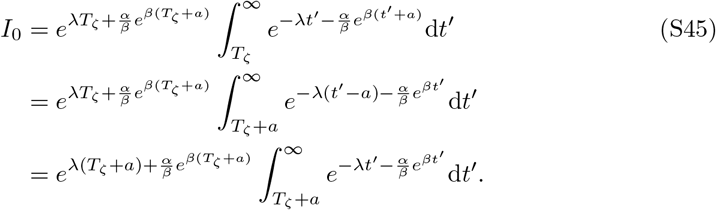

Let *v* = *e*^−*λt′*^, then d*v* = −*λe*^−*λt′*^ d*t*^*′*^ = −*λv*d*t*^*′*^. *I*_0_ gets the form:

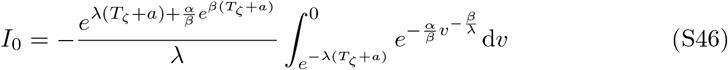

Using the change of variables 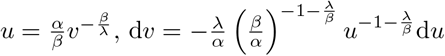, the above can be expressed as:

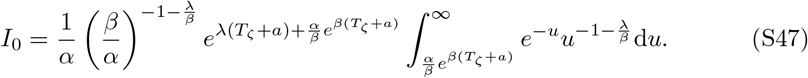

The integral to infinity in Equation S47 can be expressed as:

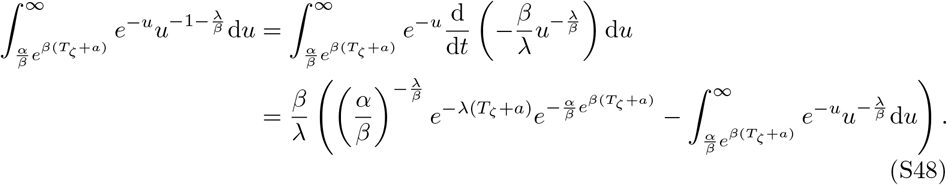

Notice that

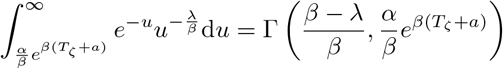

With 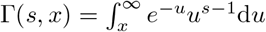. Then Equation S47 gets the form:

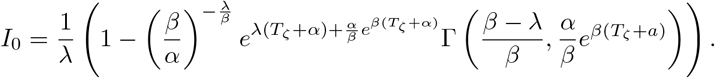

### Evaluating *I*_1_

We have:

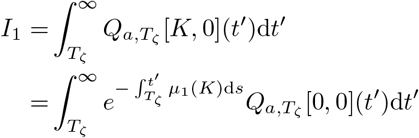

Since *µ*_1_(*K*) is a constant we get:

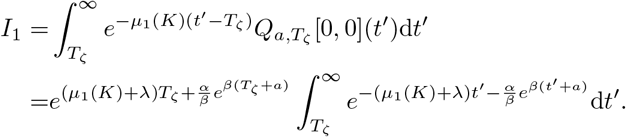

Notice that the above expression is identical to *I*_0_ (Equation S45) when replacing *λ* with *λ*^*′*^ = *λ* + *µ*_1_(*K*). We then get:

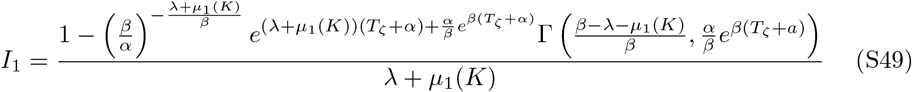

this is well defined in the case where 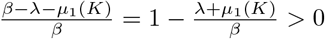. In the case where 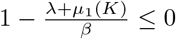, we have:

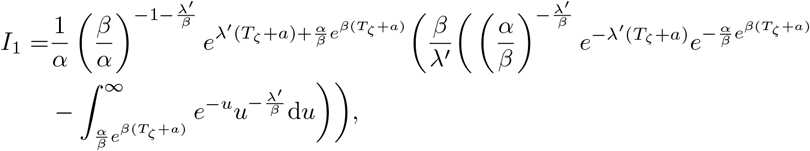

*λ*^*′*^ = *µ*_1_(*K*) + *λ*, see Equation S48 and S47. Let

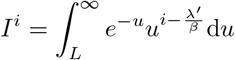

With 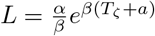. Then

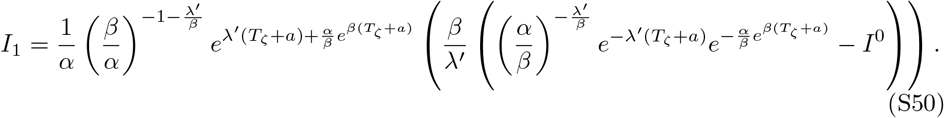

#### Theorem E.1

*If* 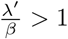 *the integral I*^0^ *is given by:*

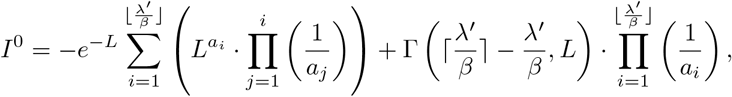

*where* 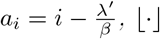 *denotes the lower integer part of* · *and* ⌈·⌉ *the upper integer part of* ·.

Let *i* be a positive integer. Then

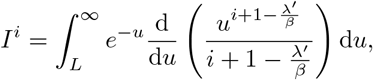

and by integrating by parts we get

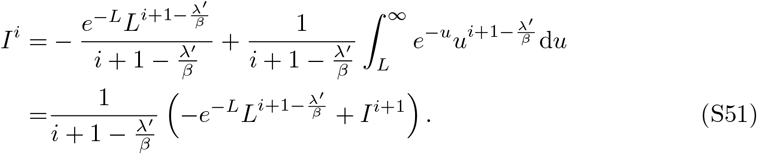

Notice that

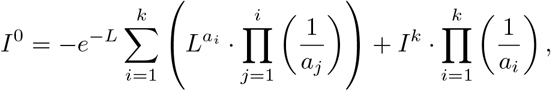

with 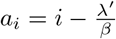. If 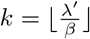 then 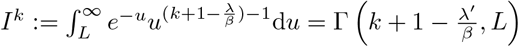 and we get:

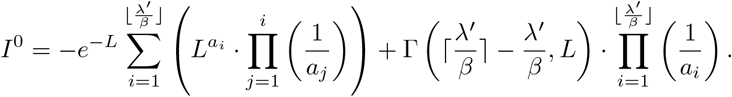

In the case where 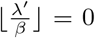, then *I*^0^ can be directly expressed in terms of a gamma function. Using Theorem E and Equation S50 we get

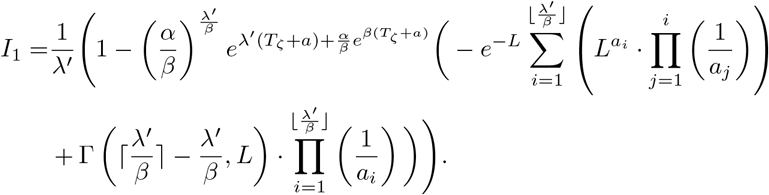

## F Expressing the CLP in Mayer form

All integral terms in Equation 21 can be expressed as terminal-state contributions, since the integrands correspond to derivatives of state variables. The final term, however, is an improper integral, as its upper bound extends to infinity. Nevertheless, this term can be rewritten in terms of *I*_0_. Indeed,

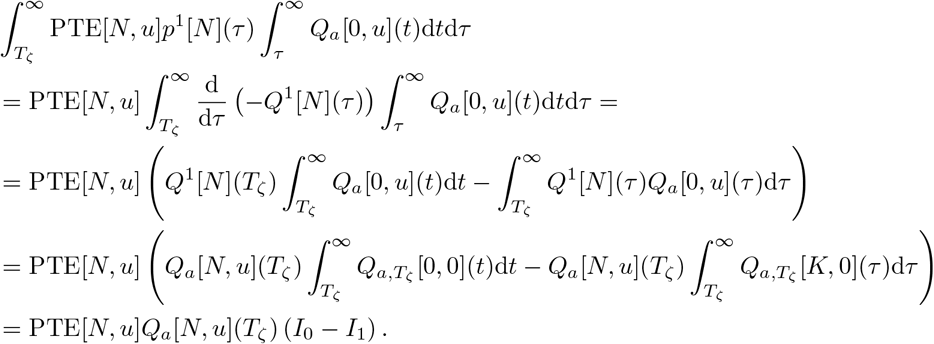

Then CLP gets the form:

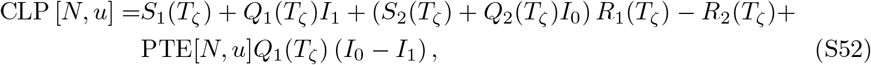

with

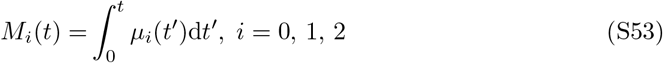

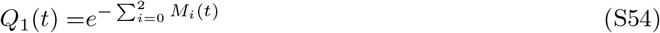

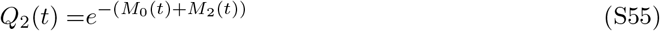

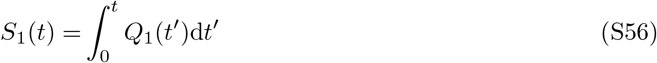

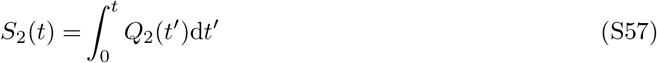

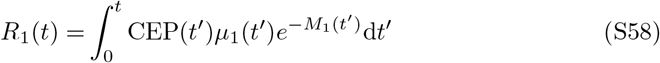

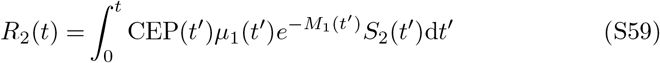

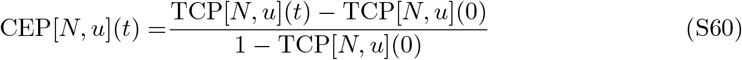

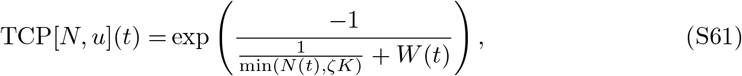

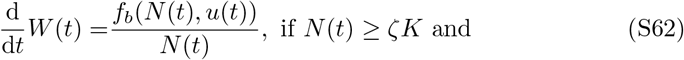

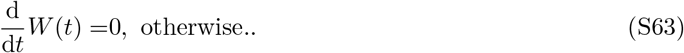

## G Optimal Control Algorithm

Here, the direct method used to maximize the CLP is presented. The control is approximated with piecewise constant functions that can change values in *p* discrete time points, *t*_1_, …, *t*_*p*_ which will be called in the following *discretisation times*. Let *t*_0_ = 0 be the starting time and *t*_*p*_ the time horizon *T*_0_. The discretisation times are in non-decreasing order and consecutive discretisation times are allowed to coincide. The control is given by

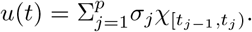

with *σ*_*j*_ ∈ [0, 1], 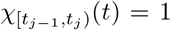 if *t* [*t*_*j*−1_, *t*_*j*_) and 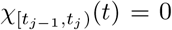 otherwise. Any piecewise constant control with at most *p* switches can be expressed by a set of discretisation times (*t*_1_, …, *t*_*p*_)^tr^. Controls with *m < p* switches can be constructed when there exist *p* − *m* couples of consecutive discretisation times that coincide (i.e. 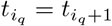 for *i*_*q*_ ∈ {1, …, *p* −1} with *q* = 1, …, *p* − *m*), since the control is fixed at the time-intervals 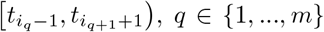, *q* ∈ {1, …, *m*}. Since no drug is administered for *t > T*_0_, we do not discretise the interval [*T*_0_, *T*]; however the state variables are evaluated until *T* using forward integration.

To accurately evaluate optimal discretisation times, we employ the time scaling method used in [46]. Let *θ*_*i*_, *i* ∈ {1, …, *p*} be positive numbers that satisfy 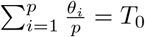.

Then each *t* ∈ [*t*_*q*−1_, *t*_*q*_), *q* = 1, .., *p* can be expressed as

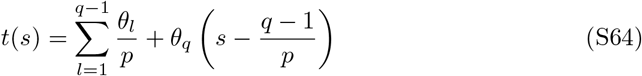

With 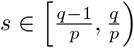. Notice that the discretisation times can be written as 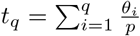 and that *t*_*p*_ = *T*_0_.

We express the state variables and the payoff as functions of 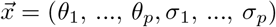, and denote the state vector with

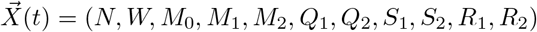

where the ODEs are expressed by the system of Equations:

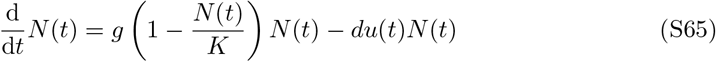

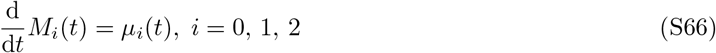

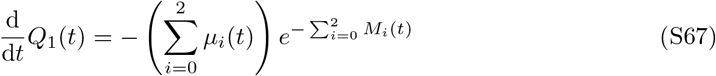

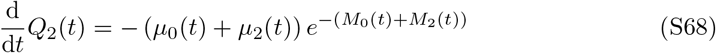

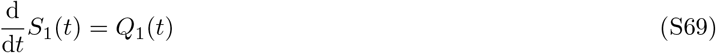

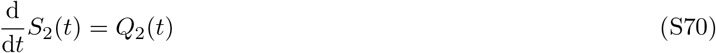

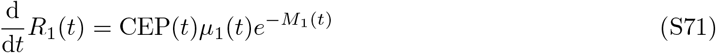

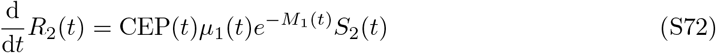

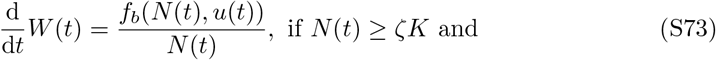

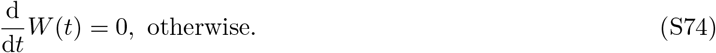

The CEP is given by Equation 6. The integrals *I*_0_ and *I*_1_ depend solely on the choice of the parameter *T* and thus are known constants (Equations 22 and 23).

We solve the ODE system for each time interval 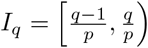:

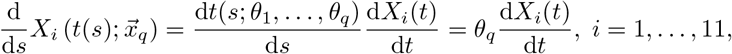

with 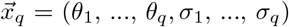 and initial conditions 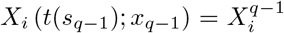 evaluated in the previous iteration (when solving for the interval *I*_*q*−1_). We evaluate the states at the saturation time *T* by solving the ODEs with no time scaling. The payoff is then expressed as a function of 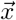

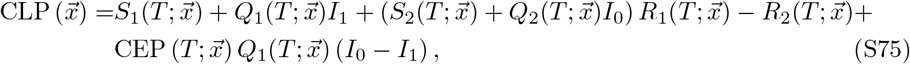

and is minimize using automatic differentiation. Computations are halted and return an error if *N*(*T*) *< ζK*.

## H Robustness of Optimal Solutions to Collocation Resolution

In Figures 7-9, the optimal control problem was solved for 64,800 different combinations of initial tumour sizes and drug tolerance intervals. To accelerate computation, the problem was initially solved using only 2 discretisation time points, restricting attention to solutions with at most one switching point. To assess the accuracy of this approximation, we also solved the optimal control problem using 30 discretisation time points for a smaller subset of parameter values. The resulting optimal solutions were of bang-bang type and matched those obtained using only 2 discretisation time points. A direct comparison between the optimal drug administration regimens computed with 30 and 2 discretisation time points is presented in Figure S20, demonstrating qualitatively similar results.

**Figure S20.**
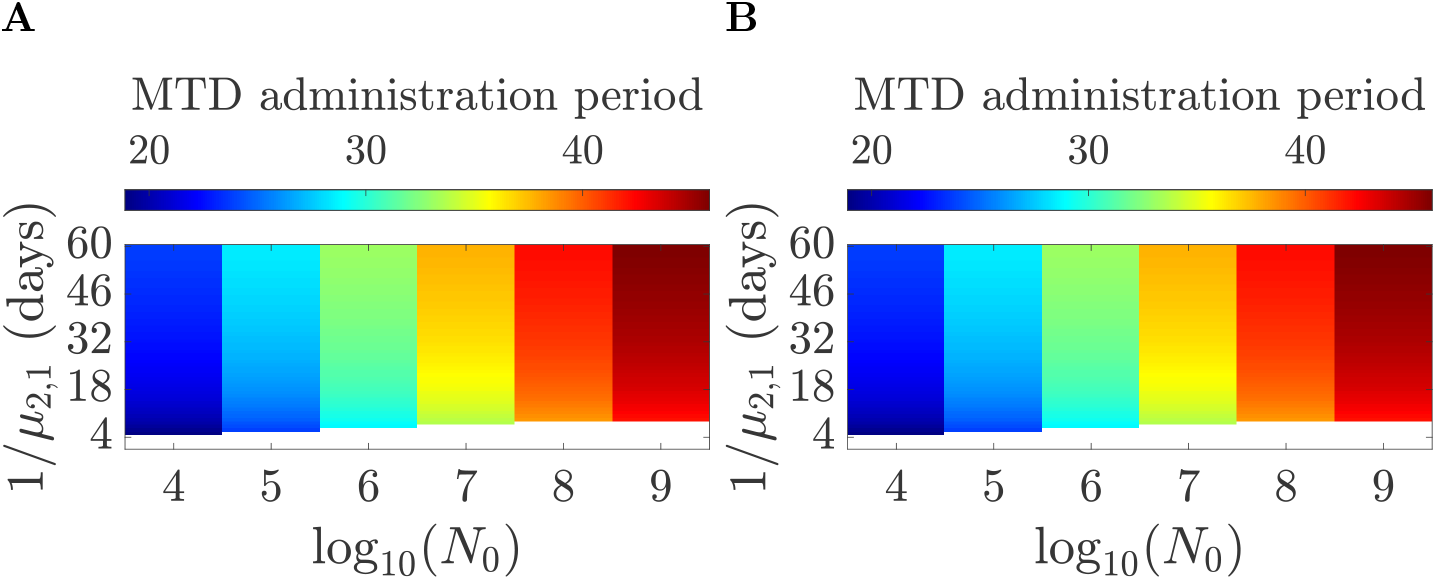
The optimal drug administration period after optimisation with **A** 30 discretisation time points and **B** 2 discretisation time points respectively. Parameters are as in Figure9.

## I Estimating the tumour and drug induced mortality rates using simulated data

In this section, we present the estimation of the parameters *µ*_1,1_ and *µ*_2,1_ using simulated data.

To generate the dataset, we considered *A* patients treated with continuous MTD for three weeks and followed for a total of 1 year. We perform the simulation based on the following: if *X* is a continuous random variable with CDF *F*_*X*_, then the random variable *U* = *F*_*X*_ (*X*) is uniformly distributed on [0, 1].

For each patient *i* = 1, …, *A*, we generated a random number *r*_*i*_ ~ *U* (0, 1). This value represents the cumulative probability of death at time *t*, that is,

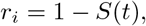

where *S*(*t*) is the survival probability defined in equation 34. Using the numerical solution for *S*(*t*) at a pre determined time grid *t*_0_ = 0, *t*_1_, …, *t*_*k*_ (*k* the number of considered time intervals (*t*_*i*−1_, *t*_*i*_]), we then determined the corresponding time interval of death (*t*_*i*−1_, *t*_*i*_], by identifying the smallest *t*_*i*_ such that *t*_*i*_ ≥ *t*.

Deaths occurring more than 1 year after the start of treatment were treated as censored observations, since no follow-up beyond this time was considered. We thus have the following events: death within the interval (*t*_*i*−1_, *t*_*i*_], *i* = 1, …, *k* and death after time *t*_*k*_. This discretization ensures that the bins can be treated as independent observations.

We denote with *n*_*i*_, *i* −1, …, *k* the number of people that died within each interval *i*, and with 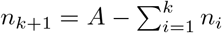 the number of people that survived longer than a year. Thus we simulate the dataset (*t*_*i*_, *n*_*i*_), *i* = 1, …, *k* + 1 and we define 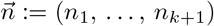.

The model predictions for *i* ≠ *k* + 1 are given by

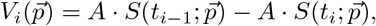

where, *S*(*t*) is the probability that the patient is alive at time *t* and 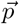 is the parameter vector. Thus, the two terms represent the expected number of individuals alive at the beginning and at the end of interval *i < k* + 1, respectively and so 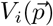 corresponds to the expected number of deaths occurring during interval *i*. For *i* = *k* + 1, we denote with 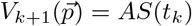 the observed number of patients that are alive a year after the start of the therapy.

The probability of observing the data, for a given set of parameter values follows a multinomial distribution, giving the likelihood:

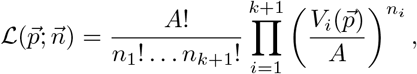

We minimize 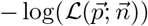 with respect to 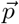. However it suffices to minimize

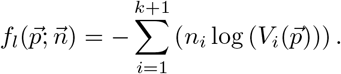

## J Correlation of the inferred parameters and of the optimal solutions based on the inferred parameters for the heterogenous and homogenous populations

In the homogenous population the correlation matrix of the inferred *µ*_1,1_, *µ*_2,1_, *g, d* and the optimal drug administration period (based on the estimated parameters) *x*_opt_ is given by

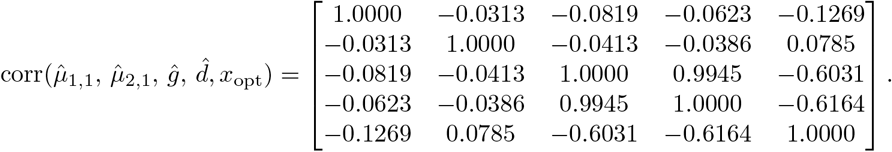

For coefficient of variation 0.1 the correlation matrix is:

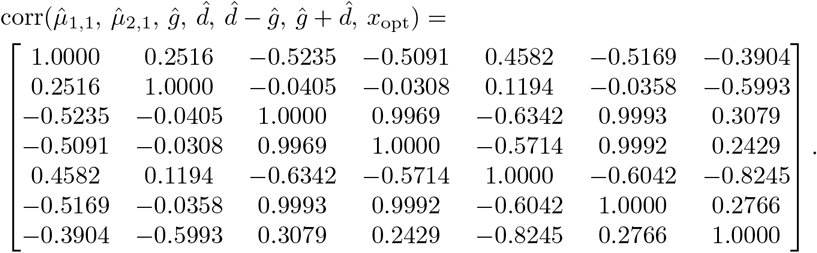

For coefficient of variation 0.25 the correlation matrix is:

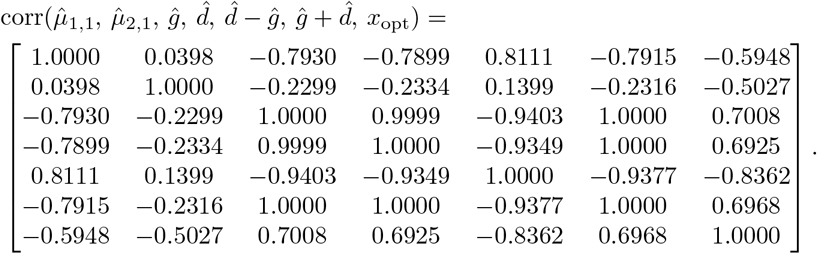

